# Causal Inference in Studies with Functional Unmasking: Psychedelics and Beyond

**DOI:** 10.64898/2025.12.05.25341713

**Authors:** Gabriel Loewinger, Mats J. Stensrud, Sandeep M. Nayak, David Yaden, Alexander W. Levis

**Affiliations:** Machine Learning Core, National Institute of Mental Health; Institute of Mathematics, École Polytechnique Fédérale de Lausanne; Center for Psychedelic and Consciousness Research, Johns Hopkins University School of Medicine; Center for Causal Inference, Department of Biostatistics, Epidemiology and Informatics, University of Pennsylvania

**Keywords:** psychedelics, psychiatry, causal inference, randomized controlled trials, semi-parametric mediation analysis

## Abstract

In clinical trials for mental health treatments, functional unmasking (unblinding) is a widespread challenge wherein participants become aware of their assigned treatment. Unmasking is especially concerning with psychedelics, due to the near unmistakable acute effects (the “trip”), resulting in uncertainty about whether outcomes following treatment reflect true therapeutic properties of the interventions, or placebo-like effects. We present a counterfactual conceptualization of unmasking that 1) formalizes the shortcomings of many existing statistical and experimental design solutions (e.g., dose-response, active controls), and 2) demonstrates how modern causal inference approaches can be applied to isolate effects devoid of this “contamination.” Our results reveal feedback mechanisms between perceived therapeutic benefits and expectancies that can render traditional methods prone to obscuring or exaggerating therapeutic benefits. Our proposal motivates trial designs and statistical methods that can be implemented to mitigate the impacts of functional unmasking.

## Introduction

Participant functional unmasking (or unblinding) frequently arises in mental health research, where acute effects of a treatment often allow participants to accurately guess their treatment assignment. This, in turn, can lead to differing “hopes” for improvement across treatment arms, possibly affecting study conclusions. At the heart of this discussion lie questions about causal effects: 1) does the compound itself *cause* therapeutic benefits, or 2) are functional unmasking and resulting discrepant hopes across treatment arms responsible for treatment effects. On the one hand, randomization ensures that average between-arm differences in the outcome are caused by treatment; the hopes, themselves, can be cast as part of the therapeutic mechanism or treatment effect. However, if hopes, or *expectancies*, strongly influence treatment effects, some may be concerned that therapeutic benefits may not be durable, or that study results may generalize poorly if these expectancies change across time or populations. We argue that introducing specific formal summaries of treatment effects (*causal estimands*) helps clarify and address the issue of unmasking.

Whether functional unmasking invalidates causal inferences from randomized clinical trials (RCTs) is a debate that is especially active in psychedelic research due partly to the FDA’s 2024 rejection of the Lykos pharmaceutical application for midoamfetamine (MDMA) as a treatment of PTSD. The FDA advisory committee’s concerns over study bias from expectancy effects and functional unmasking (Colloca and Fava, 2024) have led to a slew of publications (Colloca and Fava, 2024; Rosenbaum, 2024; Nayak and Zahid, 2025; Schenberg, 2025; Szigeti et al., 2025) and dedicated meetings (Cheung et al., 2025; Reagan-Udall Foundation for the FDA, 2024). To illustrate, a statistical consultant to Lykos presented an analysis, displayed in Figure 1A, at the FDA meeting showing that treatment effects (i.e., differences in followup PTSD levels between treatment arms) varied substantially depending on whether patients believed they had received MDMA. Similar analyses have been applied in other psychedelic RCTs to compare treatment effects across levels of post-treatment beliefs or expectancies (Bershad et al., 2019; van Elk et al., 2022; Lii et al., 2023a; Cavanna et al., 2022). Intuitively, if these analyses reveal larger effects among those who believe they received active treatment, it is tempting to conclude that placebo effects are inflating estimates of the therapeutic benefits. We appreciate that these analyses are often exploratory, but we caution that causal theory reveals that such approaches can be highly misleading due to so-called *collider bias*. In fact, we illustrate in Figures 1B-C and Appendix E.4.2 that conditioning on post-treatment variables in this manner (i.e., stratifying analyses on these variables or including them as covariates in regression models without formal causal estimation tools) can obscure treatment effects so strongly that highly beneficial treatments wrongfully appear as if they were harmful. This example, in addition to other common pitfalls that we have observed in the literature, highlight that formal causal theory is needed to correctly estimate the desired causal effects in studies with functional unmasking.

**Figure 1.**
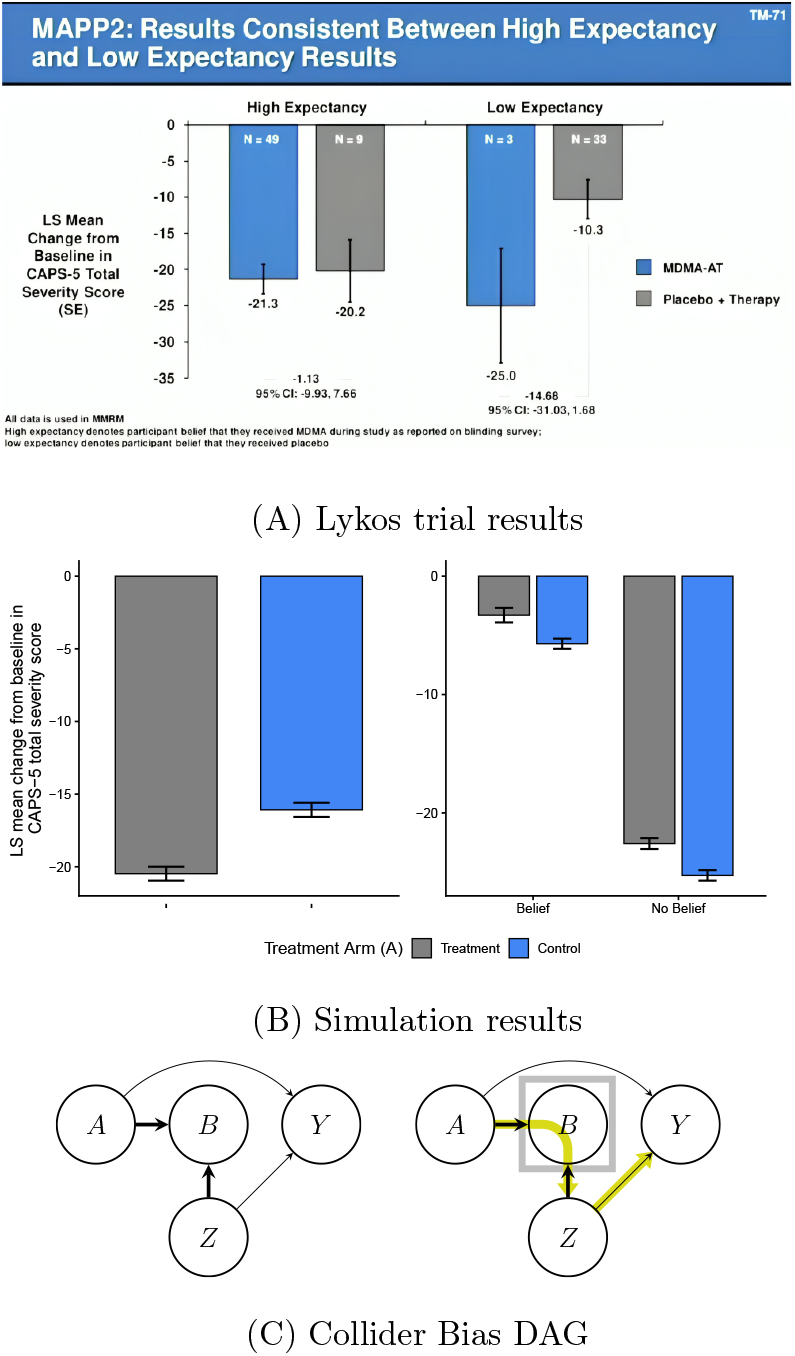
Bias from analyzing results stratified on post-treatment variables. (A) Slide presented by a statistical consultant to Lykos at the FDA Advisory meeting hosted on June 4, 2024. (Source: https://psychedelicalpha.com/news/live-coverage-fda-advisory-committee-reviews-mdma-assisted-therapy-for-ptsd). This figure was also reproduced in Colloca and Fava (2024) who note that “Presented is LS [Least Squares] mean change in the clinician-administered PTSD scale for DSM-5 (CAPS-5) total severity score.” (B) Simulation results to illustrate bias from stratifying on post-treatment variables (e.g., belief). Error bars indicate 95% confidence intervals. [Left] We compare the change from baseline PTSD scores (as shown in (A)) across treatment arms. There is a strong PTSD reduction from treatment, showing that the treatment was simulated to be beneficial; concretely, the controlled direct effect (as introduced later in the paper) of treatment, fixing belief, was negative (beneficial) for all subjects. [Right] The within-belief level differences, which wrongly makes it appear as if the treatment is instead harmful (i.e., the treatment effect sign is opposite to that of the left panel). The discrepancy between the two figures illustrates how stratifying on post-treatment belief can lead to such strong bias that the analysis suggests the opposite conclusion: the data are simulated with a large beneficial effect of treatment (negative) but the stratified analysis makes it wrongly appear as if the treatment effect is harmful (positive). This is an example of collider bias. We detail this simulation setting in Appendix E.4.2. (C) Directed Acyclic Graphs (DAGs) to illustrate collider bias in comparison of treatment effect estimates across stratum of post-treatment beliefs/expectancy. We use the notation *A* → *B* to express that treatment, *A*, is a cause of post-treatment belief, *B*. In this DAG, we denote *Z* to be common causes of belief *B* and the outcome variable *Y* (e.g., pre-treatment expectancies). The DAG on the left illustrates how an analysis that omits post-treatment belief *B* (e.g., comparing the mean outcome across treatment arms) is valid: post-treatment variables *B* are *colliders* on the *A* → *B* ← *Z* path (represented by the thick arrows). This *blocks* confounders, *Z*, of the *B* → *Y* pathway, from exerting causal effects mediated through the *A* → *Y* pathway (i.e., the path from *A* → *B* ← *Z* → *Y* is blocked). The DAG on the right illustrates why stratifying treatment effect estimates on *B* (or including *B* in a regression without formal causal estimation methods) invalidates this analysis: stratification on *B* (represented by the box) *opens* the pathway from *A* → *B* ← *Z* → *Y*, thereby introducing bias from *Z* and *B*. Thus conditioning on *B* in this manner could lead to a beneficial treatment appear (incorrectly) harmful.

Carefully applying causal tools can allow recovery of treatment effect estimates in psychedelic trials comparable to those that would be obtained in a successfully masked study. In other domains, applications of tailored causal methods to observational data have yielded treatment effect estimates comparable to those from subsequent RCTs. Causal inference methods have proved critical for demonstrating treatment efficacy in both high-profile RCTs and observational studies of, for example, HIV (Robins, 1987, 1986), cancer (Dickerman et al., 2019), hormone therapy in women (Hernán et al., 2008), and COVID-19 (Dickerman et al., 2022). Together, the widespread applications and successes of causal inference have resulted in the awarding of the Turing award in 2011 (Pearl, 2011), Nobel prize in 2021 (Organization, 2021), and Rousseeuw Prize for Statistics in 2022 (King Baudouin Foundation, 2022b,a). These successes were enabled in part by modern causal estimation methods based on semiparametric theory that can incorporate powerful machine learning algorithms. These tools have not, however, been widely adopted to combat methodological challenges in mental health research.

We aim to apply *counterfactual* and *graphical* causal theory to 1) argue that statistical approaches and RCT designs (e.g., dose-response, active controls/comparators, treatment under anesthesia) applied in the psychedelic literature will not alone resolve the unmasking challenge, 2) propose modern causal inference methods to account for unmasking in existing RCT designs, and 3) suggest new designs that can mitigate and/or quantify the effects of unmasking. As a solution to a challenging instance of functional unmasking, our work can be adapted to studies of treatments beyond psychedelics.

### Expectancy: Nuisance or Mechanism?

To introduce our proposal, we first define functional (un)masking in formal causal language, illustrated by an example hypothetical RCT studying treatment for depression. A sample of participants are randomized at baseline to receive a single dose of psilocybin, or methylphenidate as an active comparator/control. Following standard causal inference notation (Hernan and Robins, 2024), we denote the treatment (psilocybin) arm as *A* = 1 and the control (methylphenidate) arm as *A* = 0. To focus on the main arguments, we assume no dropout and full adherence to the assigned treatment. We measure baseline (pre-treatment) covariates, *X*, like “hype” (pre-treatment hopes), age, and sex. Six months after treatment, we measure follow-up depression levels, *Y*, and compare the averages of this outcome variable across arms to estimate the *average treatment effect* (ATE). Shortly after treatment, we also measure: 1) *beliefs, B*, indicating which treatment a participant thinks they received; 2) post-treatment *expectancies, E*, indicating whether they think their depression *Y* will improve/worsen (see Appendix A for a comparison of these definitions with previous work); as well as 3) a collection of variables (denoted by *Z*) that affect post-treatment expectancy and the outcome. The vector *Z* might include, for example, the experience of the trip. To avoid confusion, we use *hype* to refer to pre-treatment hopes, and *expectancy* (*E*) to refer to post-treatment hopes. To introduce counterfactual causal reasoning, we define the *counterfactual* (or *potential outcome*) *Y* (*a*) which denotes the outcome (depression levels) that *would be* observed if, potentially counter-to-fact, an individual received treatment level *a*. We use uppercase *A* to denote the actual observed treatment value, and lowercase *a* for the intervention value that we set the treatment to. This frames causal inference in terms of a hypothetical setting where we could observe follow-up depression levels of a given participant in two worlds: one in which they received psilocybin and one in which they received control. Although we cannot observe data from both worlds for the same individual, we can use real data to estimate average outcomes under each treatment condition. For example, in our psilocybin RCT, we can estimate the average of each counterfactual as the mean outcome in the respective treatment arm. More generally, our strategy for targeting more complex causal questions proceeds through (i) articulation of the research question of interest, formalized as a causal estimand, and (ii) delineating when and how to estimate it using data from new or existing experiments.

We use this framework to argue that resolving the functional unmasking debate boils down to carefully accounting for (post-treatment) expectancy, *E*, statistically or experimentally. The expectancy/belief distinction is critical because it clarifies why treatment effect estimates can be “contaminated” even when masking is successful. In fact, to those skeptical of psychedelic’s therapeutic benefits, randomized open-label (unmasked) studies without expectancy effects may be less worrisome than successfully masked studies where the treatment/control cause differing expectancy levels. For example, consider a randomized open-label study comparing participants’ absorption levels of two Vitamin E oral supplement formulations that have equivalent perceptible effects (e.g., side effects). If neither formulation is expected to be superior, knowledge of which one a participant has received is unlikely to systematically favor one treatment. We call this study *belief unblinded*, as participants can determine which arm they were assigned to, and *expectancy blinded*, as each participant in this study is assumed to have the same expectancy levels regardless of treatment assignment. On the other hand, *belief blinded* – *expectancy unblinded* RCTs appear more concerning. In such studies, treatments cause differences in expectancy levels across arms even though participants cannot determine which treatment they received. For instance, consider the (potentially unrealistic) depression treatment study where salvia is used as an active control/comparator for smoke-able DMT among a psychedelic-naive sample. Suppose salvia’s psychoactive effects lead to belief blinding, but its dysphoric properties lead to negative expectancies about how treatment will influence depression levels (i.e., expectancy unblinding). Then, even if neither DMT nor salvia possessed lasting therapeutic benefits, the outcomes of this hypothetical RCT may suggest DMT is beneficial only because salvia led to more negative expectancy levels than DMT.

To analyze these ideas more precisely, we use the variable definitions from our hypothetical psilocybin RCT and encode their relation in a directed acyclic graph (DAG) (Hernan and Robins, 2024) in Figure 2A. In the DAG, the notation *A* → *B* expresses that treatment, *A*, is a cause of belief, *B*. The DAG shows how all past variables are potential causes of all future variables, except *A*: treatment has no causes because of randomization (no arrows point to *A*). Thus, baseline variables *X*, like hype, might modify the effects of *A* on *Y* but do not *confound* the *A* → *Y* pathway (even in the presence of functional unmasking) because *X* is not a *common cause* of these variables.

**Figure 2.**
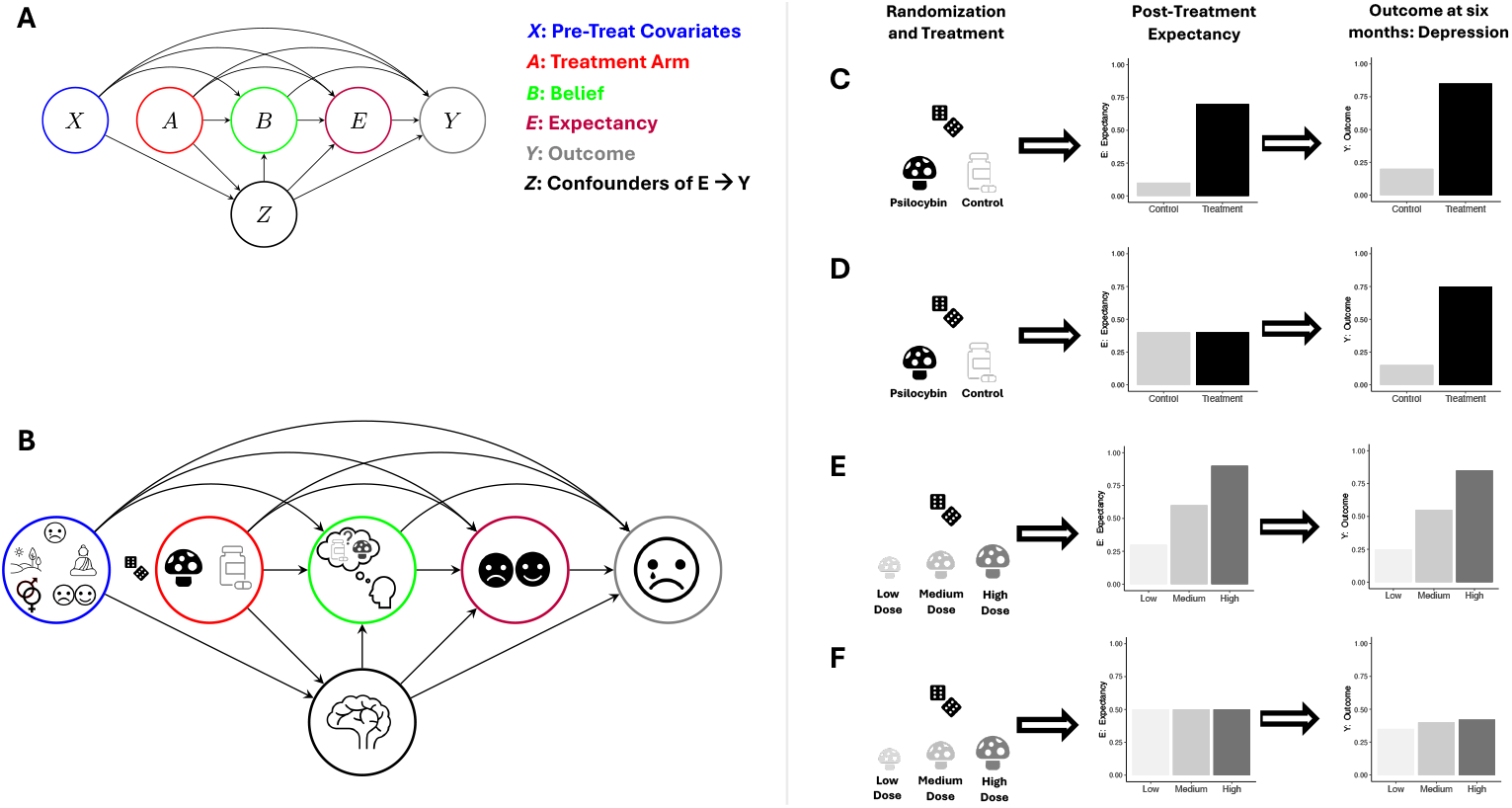
Causal conceptualization of psychedelic trials in the presence of functional unmasking. (A) Causal directed acyclic graph (DAG) (Hernan and Robins, 2024) the hypothetical psilocybin RCT with variables: *X* – baseline (pre-treatment) covariates; *A* – treatment (*A* = 1) control (*A* = 0) arm; *B* – participant belief that they received treatment (*B* = 1) or control (*B* = 0); *Y* – outcome variable (e.g., depression); *E* – expectancy about how they think their outcome will change from treatment; *Z* – post-treatment confounders of *E* → *Y* path (i.e., post-treatment common causes of both *E* and *Y*). The direction of *B* → *E* instead of *E B* may be switched depending on the study design and source of unmasking. This DAG allows one to define belief-, expectancy-, and complete-blinding in terms of causal pathways (see Appendix Table 1). (B) The same DAG as in A but with graphical illustration of example variables to include. *X* contains pre-treatment depression, set and setting, biological sex, and hype (pre-treatment hopes/attitudes). *Z* contains the acute effects of the trip (represented by the brain). *A* has a dice icon next to its node to indicate no arrows point into it because of randomization (i.e., there are no causes of *A*). Post-treatment expectancy *E* is indicated by smiling/frowning face. *Y* is post-treatment depression. (C) Sequence of 1) randomization/treatment, 2) resulting expectancy changes, and 3) the outcome. This sequence illustrates how active (*A* = 1) and control (*A* = 0) treatments often result in changes in both expectancy and the outcome leading to the possibility that expectancy *mediates* treatment effects. (D) Illustration of the idealized setting where even when (post-treatment) expectancy is experimentally “set” to be the same value for a given individual regardless of treatment/control arm assignment, the treatment effect (differences in the outcome across arms) remains pronounced. This illustrates a setting where expectancy does *not* mediate treatment effects substantially. However, if expectancy *does* mediate these effects, then other approaches are needed to account for expectancy effects. For example, (E)-(F) illustrates why dose-response studies do not necessarily resolve the functional unmasking problem: expectancy may itself exhibit a dose-response relationship with the treatment. Thus, the dose-response effect in the outcome may be mediated by this expectancy dose-response profile. (F) shows what might be observed in practice (without experimentally controlling expectancy). (G) shows what would happen in a psychedelic dose-response study if therapeutic effects are in fact entirely mediated through expectancy: we experimentally “set” expectancy to be the same across treatment doses (within an individual) and the treatment effects on *Y* disappear because they were almost entirely mediated through the dose-response expectancy relationship.

These DAG paths distill complex mechanisms without discarding critical information. For example, the *A* → *Y* or *A* → *B* pathways may be mediated by side-effects, which relate to benign/malicious unmasking (Howick, 2011), as discussed below. Similarly, the effect of expectancy on depression (*E* → *Y* path) may be mediated by behavior change (e.g., diet). There may also exist biological/psychological mechanisms (i.e., causal pathways) that are not mediated through *E* (e.g., represented by the direct *A* → *Y* path), but are critical to psilocybin’s effectiveness. Thus, although simplified, a more extensive DAG that includes these intermediate mechanisms does not, in most cases, change the causal structure necessary to identify how to mitigate the impacts of functional unmasking.

**Table 1:** Pre-efficacy blinding types. We use the notation *A* ↛ *B* to indicate that there are no paths from *A* to *B*. Belief blinding (*A* ↛ *B*) means that the treatment does not cause belief (i.e., differences between-arms in the treatment that participants believed they received). Conversely, expectancy blinding (*A* ↛ *E*) means that the treatment does not cause post-treatment expectancy (i.e., differences between-arms in expectancies about how their outcome will change depending as a result of their (perceived) treatment). Complete blinding indicates that both of these blinding types are achieved. Belief and expectancy blinding often occur together, but the examples in the main text Section illustrate cases where a study might achieve one type but not the other. We describe preefficacy blinding types here but these can be generalized to post-efficacy blinding types with similar notation.

|  |  |
| --- | --- |
| Belief Blinding | $A \nrightarrow B$ |
| Expectancy Blinding | $A \nrightarrow E$ |
| Complete Blinding | $A \nrightarrow B$, and $A \nrightarrow E$ |

Whether treatment effects are or are not mediated through *E* may seem arbitrary to someone who views expectancy effects as a key mechanism of psychedelics. We argue, however, through the formal approach we propose, that understanding the magnitude/durability of the causal effects of treatment and (post-treatment) expectancy together can directly probe the concerns of critics (e.g., Muthukumaraswamy et al. (2021); Butler et al. (2022)) that unmasking is unfairly driving the large effect sizes reported. For instance, a skeptic that views expectancy as a *nuisance* might only be convinced of psychedelics’ therapeutic potential if treatment effects are shown to be large in a study where *E* was effectively fixed to be high (“hopeful”) or low (“not hopeful”). Conversely, those who conceptualize *E* as a legitimate *mechanistic* component of the treatment, may seek to demonstrate that the independent therapeutic benefits of expectancy are durable enough to be considered a reliable therapeutic effect. Thus, for both the nuisance and mechanistic camps, estimating the joint effects of treatment and expectancy lies at the foundation of the unmasking debate.

### A Counterfactual Description of Functional Unmasking

Our counterfactual framing clarifies *why* accounting for expectancy mitigates the downsides of functional unmasking, *what* quantitative summaries of treatment effects (causal estimands) isolate these unwanted expectancy effects, and *how* to estimate them in RCTs. We start with the *why*. Our approach reveals that expectancy is a causal mediation problem, not a confounding problem: since unmasking is a post-treatment phenomenon, *E* and *B* are intermediate variables between *A* and *Y*, not common causes (confounders) of *A* and *Y* (see Figure 2 for intuition). This conflicts with what we believe is a common misconception that unmasking *leads to* confounding. For instance, Muthukumaraswamy et al. (2021) state that “[p]sychedelic RCTs are likely confounded by de-blinding” and if the RCTs “cannot be ade-quately masked…[they] will always be confounded.” We argue that “confounding” of the treatment–outcome relationship will not occur in RCTs from unmasking because randomization ensures there exists no common causes of the treatment and outcome (i.e., no confounding). Thus, RCTs enable valid estimation of the ATE even under unmasking. That ATE may, however, be partially *mediated* through unwanted (post-treatment) expectancy effects. This emphasizes why accounting for hype (pre-treatment expectations) may not, on its own, fully address the unmasking challenge. For example, Aday et al. (2022) write that “without effective condition masking, it is virtually impossible to maintain the independence of the […] [treatment], as it is confounded by participant [pre-treatment] expectations.” But *A* is randomized so hype cannot *confound* the *A* → *Y* pathway. In fact, hype will be balanced on average across arms. Thus, statistically/experimentally controlling for hype will not alone resolve the unmasking challenge. Hype *is* likely critical to incorporate into analyses, but as a confounder of the *E* → *Y* relationship. For example, unmasking may “unleash”/“unlock” pre-treatment hopes/attitudes to strongly influence (post-treatment) expectancy and depression.

Second, by conceptualizing treatment effects as mediated through expectancy, our framing provides formal statistical methods and RCT designs to isolate study effects of treatment that control expectancy (the *what*). Specifically, the *controlled direct effect* (CDE) causal estimand isolates this effect. We discuss the formal definition below, but conceptually, the CDE quantifies the average between-arm difference in *Y*, in a hypothetical study where we experimentally *set* (post-treatment) expectancy levels to the same value across arms. Formally defining the causal effects that we target is critical: while others have appealed to DAGs to describe unmasking in psychedelic RCTs, we believe that these proposals have drawbacks. In fact, as discussed below, some of these methodological proposals (e.g., Szigeti and Heifets (2024)) can result in estimates that are so misleading, they can make beneficial treatments appear harmful, or vice-versa (see Appendix E.4.1 for a detailed derivation).

Third, the DAG clarifies that we can estimate the CDE if the correct variables are measured, or if certain RCT designs are implemented (the *how*). Specifically, in RCTs that only randomize *A*, one must measure and adjust for all common causes (*X* and *Z*) of expectancy *E* and depression levels *Y*. We discuss what these variables might be below (e.g., *X* should include hype). Unlike past proposals in the psychedelic literature, we propose application of formal and modern causal inference methodology. Importantly, estimation differs from the intuitive, but invalid, strategy of *conditioning* or *stratifying* on a measurement of expectancy. The DAG also motivates that one can instead experimentally manipulate *E* with specific types of RCT designs (discussed below). Thus these statistical and experimental strategies allow one to isolate treatment effects devoid of unwanted expectancy “contamination.”

### Classical RCTs do not always target the right causal effect

Before discussing estimands to account for unmasking, we describe drawbacks of the ATE. Because expectancy-mediated effects are often unwanted, the ATE does not always summarize the effects of interest. Conceptually, this is because the average *total effect* of *A* → *Y* may be composed of a *direct effect* of *A* → *Y* and *indirect effect* mediated through *E*: *A* → *E* → *Y*. Thus the indirect effects mediated through expectancy may exaggerate or obscure the direct (biological/psychological) effects of psychedelics that are not expectancy-mediated. In fact, treatment can induce expectancy feedback mechanisms that further complicate the interpretation of total effects.

We illustrate this concept via several RCTs in Figure 3. This phenomenon, which we term *efficacy-expectancy feedback*, describes how perceived post-treatment efficacy of the treatment affects (post-treatment) expectancy, in turn impacting the outcome itself. In some cases, this creates a “snow-balling” whereby perceived treatment benefits cause greater expectancy, feeding back to a greater perception of treatment efficacy (see the *Aspirin* example in Figure 3). Efficacy–expectancy feedback (formalized in Appendix D.1) can be cast as a special case of treatment-confounder feedback, a well-known causal inference concept (Robins, 1986; Hernan and Robins, 2024), whereby feedback mechanisms can also obscure treatment effects (Levis et al., 2024).

**Figure 3.**
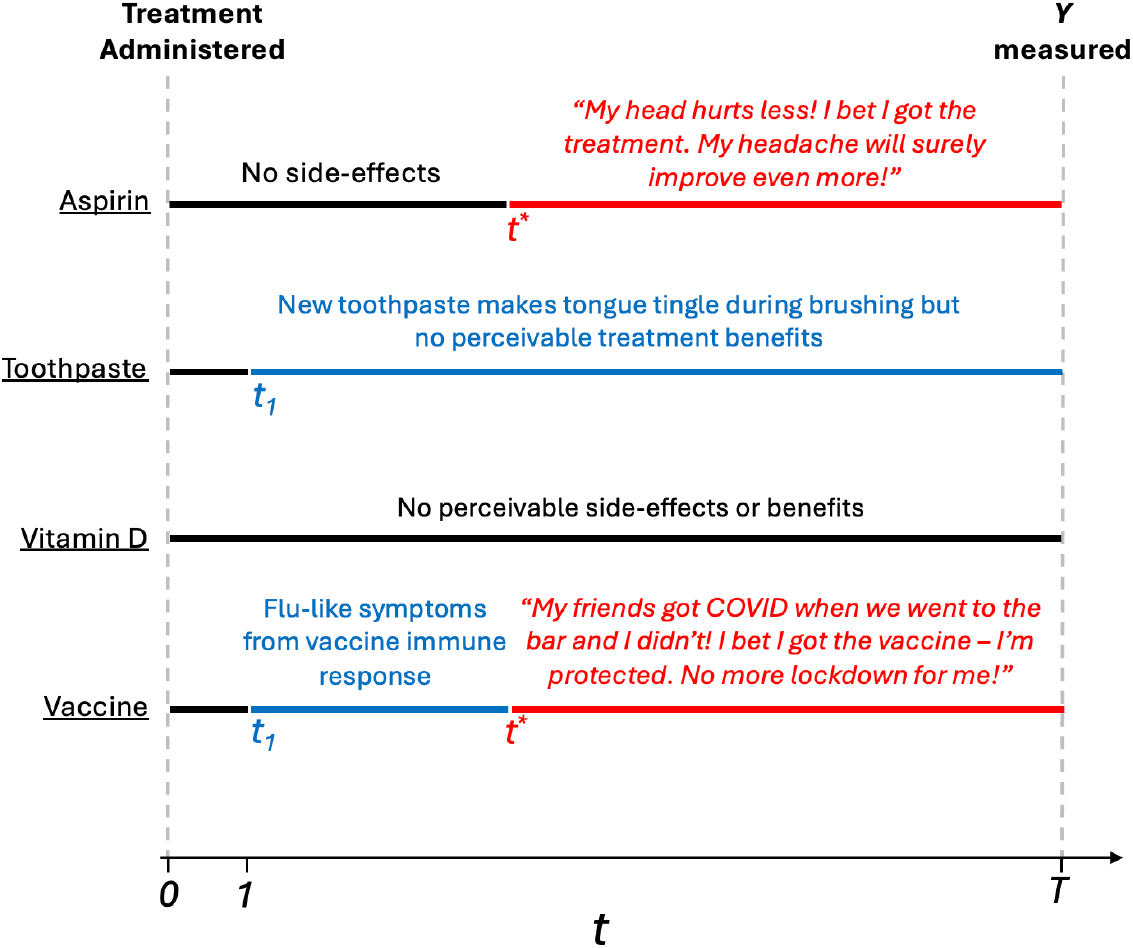
Four example RCTs with and without side-effects and/or efficacy-expectancy feedback. In all examples, the (horizontal) black line indicates the time-interval between treatment administration (*t* = 0) and the first perceivable effects (either *t*_1_ or *t*^∗^). Here, *t*_1_ is the time when perceivable side-effects occur. *t*^∗^ is the time when perceivable therapeutic benefits occur. *T* is study follow-up. The blue line indicates intervals from *t*_1_ until either *t*^∗^ or *T*. The red line indicates time-windows between *t*^∗^ and *T*. The lack of a red line means the treatment has no perceiveable therapeutic benefits. The lack of a blue line means the treatment has no perceiveable side-effects. <u>Aspirin</u> We run a double blind RCT to study the efficacy of aspirin, relative to an inactive control, to treat headaches. *Aspirin* is effectively masked and has no perceivable effects until *t*^∗^, the timepoint, aspirin (*A* = 1) arm participants begin to feel improvements in their headache. When aspirin arm participants begin experiencing a “real” treatment-induced reduction in headaches, a “snow-balling” effect occurs: the therapeutic benefit leads to greater expectancy. These patients think “my headache is improving, I bet I got the active treatment, and my headache will improve even more.” This greater expectancy further improves the perceived efficacy of the treatment, which again feeds back leading to even greater expectancy levels. This describes *efficacy-expectancy feedback*. The <u>Toothpaste</u> RCT compares two toothpastes for gum health at six month follow-up. The *A* = 1 arm toothpaste has side-effects (e.g. the tongue “tingles”) (starting at *t*_1_ = 1) that the comparator toothpaste (*A* = 0) does not. However, treatment benefits to gum health are not perceptible during the study and so there is no efficacy-expectancy feedback. The <u>Vitamin D</u> Supplement RCT compares two formulations. Neither treatment have any perceivable effects. The *COVID-19 Vaccine* induces flu-like symptoms the day after administration because of the acute immune response. Six weeks after administration, treatment arm (*A* = 1) participants notice that they did not develop COVID symptoms when exposed, but their unvaccinated friends got infected. As a result of their perceived protection, participants begin taking more risks (e.g. going to restaurants more often), inducing a form of efficacy-expectancy feedback. This increased exposure to infected individuals lead to greater infection, masking the vaccine-induced immunity. Appendix Table 2 describes the blue and red time-intervals in terms of a 2 × 2 table of side-effect timing vs. efficacy-expectancy feedback.

The hypothetical RCTs in Figure 3 also illustrate important time-windows and differentiate between 1) unmasking from perceivable side-effects versus from therapeutic benefits, and 2) expectancies before/after perceived therapeutic benefits (“efficacy”). We argue that this preefficacy/post-efficacy distinction is preferable to the benign/malicious unmasking dichotomy, defined as unmasking from perceived efficacy, or side-effects, respectively (Howick, 2011). Indeed, unmasking due to perceived treatment efficacy is often considered “benign”, but it can also exaggerate/shroud therapeutic benefits. In Appendix D, we extend the DAG in Figure 2A with longitudinal measurements of *E* and *B* to formalize these critical time-windows, and distinct types of post-treatment expectancy effects. In Figure 4, we illustrate an example of efficacy-expectancy feedback in a mathematical simulation of an RCT in which the immunity from a vaccine contributes to higher risk-taking behavior than control arm participants, even when participants are effectively belief-blinded. The higher risk-taking behavior, an example of expectancy unblinding, counteracts the vaccine-induced protection, ultimately obscuring the protectiveness of the vaccine when quantified with an ATE. However, a CDE estimate reveals the underlying vaccine effectiveness: vaccinated participants would exhibit far lower incidence than controls if we had intervened (e.g., with education) to ensure subjects in both arms had identical risk behavior (i.e., expectancy levels).

**Table 2:**
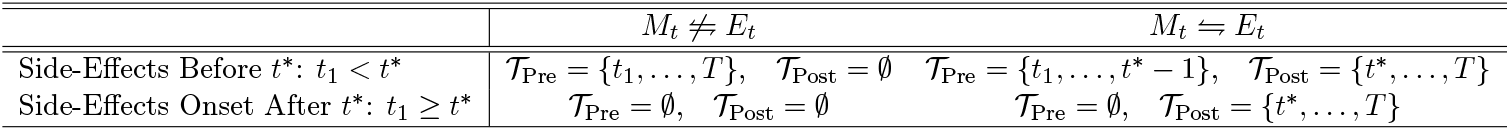
Critical time windows for effect isolation in terms of side-effect timing vs. Efficacy-Expectancy Feedback. We use the graphical shorthand, *M*_*t*_ ⇋ *E*_*t*_ to denote this feedback process, with the understanding that the elaborated graphical model is acyclic. The second row includes the case where side-effects begin after efficacy-expectancy feedback onset, *t*^∗^ (*t*_1_ ≥ *t*^∗^); the second row also applies to the special case where there are no side-effects at any point in the study. We provide example RCTs in Figure 8.

| | $M_t \not\Leftarrow E_t$ | $M_t \Leftarrow E_t$ |
| --- | --- | --- |
| Side-Effects Before $t^*$: $t_1 < t^*$ | $\mathcal{T}_{\text{Pre}} = \{t_1, \dots, T\}, \mathcal{T}_{\text{Post}} = \emptyset$ | $\mathcal{T}_{\text{Pre}} = \{t_1, \dots, t^* - 1\}, \mathcal{T}_{\text{Post}} = \{t^*, \dots, T\}$ |
| Side-Effects Onset After $t^*$: $t_1 \geq t^*$ | $\mathcal{T}_{\text{Pre}} = \emptyset, \mathcal{T}_{\text{Post}} = \emptyset$ | $\mathcal{T}_{\text{Pre}} = \emptyset, \mathcal{T}_{\text{Post}} = \{t^*, \dots, T\}$ |

**Figure 4.**
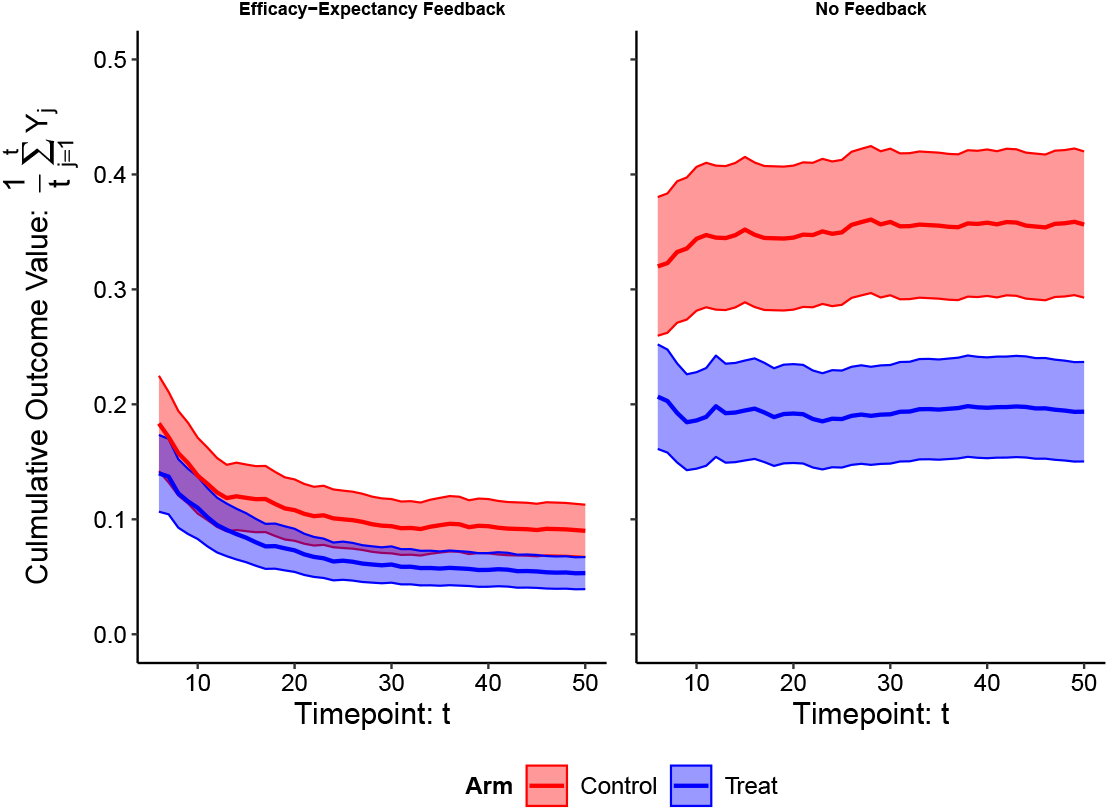
Efficacy–Expectancy Feedback Example. Vaccine RCT simulation example. [Left] Simulation results *with* efficacy-expectancy feedback: participants reduce their sexual risk behavior if they become infected. The outcome *Y*_*j*_ = 1 if a participant became infected at timepoint *j* and *Y*_*j*_ = 0 if they did not become infected. We show the average culmulative number of infections on the y-axis and timepoint on the x-axis. [Right] Simulation results from an identical simulation but *without* efficacy-expectancy feedback: participant infection throughout the study does *not* alter their risk behavior. When participants have the same risk behavior in both arms: the vaccine reduces the risk of infection for each exposure by 50%. This simulation illustrates a vaccine RCT for the sexually-transmitted infection Chlamydia among freshman college students on a campus with an outbreak. All freshman students are randomized to receive either the vaccine (*A* = 1) or a control (*A* = 0) shot before the first semester. There are no side effects and successful belief-blinding is achieved throughout. Participant infection status is tested weekly, and infected participants are given an antibiotic to treat the infection. For simplicity, we imagine the antibiotic works instantaneously, and does not prevent future infections. At the end of the study, the average number of infections each participant experienced is compared across arms. Efficacy-expectancy feedback can obscure strong treatment effects: testing positive for Chlamydia scares students into reducing their tendency to have unprotected sex. The tests therefore provide “feedback” in a manner that can alter behavior, even if participants are unsure of which treatment arm they are in. Since the data is simulated under a scenario where the vaccine is effective, *A* = 1 students become infected with a lower probability for each exposure, and so over time, they exhibit riskier behavior than controls on average (i.e., vaccine participants are “scared-off” from future unprotected sex less than controls). We provide simulation details in Appendix D.2.

This concept and the above example are relevant to psychedelic RCTs. First, belief-blinding is not sufficient to prevent expectancy feedback. Even when between-arm differences in expectancy levels arise solely from treatment efficacy (e.g., higher risk behavior resulting from vaccine protectiveness), treatment effects can still be obscured or exaggerated. Thus, even if functional unmasking in psychedelic RCTs resulted solely from a recognition of the therapeutic benefits from the trip (i.e., no placebo effects), controlling expectancy effects would still be desirable for both nuisance and mechanistic camps. Finally, this shows how careful causal analyses can reveal both expectancy feedback and therapeutic benefits that would otherwise be obscured if summarized with an ATE.

### Flexible Causal Analysis

Some of the major successes of causal inference in high-profile biomedical studies were enabled by causal methods to resolve feedback mechanisms (Robins, 1986; Hernan and Robins, 2024) akin to efficacy-expectancy feedback, highlighting the potential promise of our proposed strategy to use these tools to isolate the effect of expectancy. Specifically, we can formalize the unmasking challenge as a causal analysis of questions like “what is the difference in depression levels at follow-up among those treated and those not treated with psilocybin, in a world where we set expectancy levels to the same value across arms?” This describes, at an individual participant-level, what the causal effect of psilocybin levels are (on depression) for a fixed level of expectancy. The average of these individual-level differences is the *controlled direct effect* (Robins and Greenland, 1992), defined by the contrast of counterfactual means,

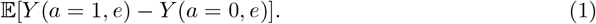

The CDE represents a contrast of effects of two joint interventions: one that sets *A* to active treatment (*A* = 1) and expectancy to *E* = *e*, and another that sets *A* to control (*A* = 0) and expectancy to *E* = *e* (illustrated in Figure 6A). The CDE can be conceptualized by an idealized *target trial*: a hypothetical experiment that naturally enables estimation of the causal effect that we seek to estimate by jointly intervening on the variables of interest (i.e., both the treatment and expectancy levels). For example, imagine the 2 × 2 design of (psilocybin vs. methylphenidate) × (low vs. high expectancy), where experimentally setting expectancy exactly to a low/high level was feasible with, for example, a consultation with a trusted physician that effectively convinces participants to be hopeful or neutral. With such a design, one could compare average depression levels *Y* across psilocybin/methylphenidate sub-arms, within the low- or high-expectancy arm. This would allow one to experimentally test for the effect of psilocybin while “controlling” for expectancy effects. This 2 × 2 design illustrates how a hypothetical expectancy manipulation is *theoretically* well-defined, even if perfect manipulation is not practically feasible (or ethical) at the time of the trial. Critically, the CDE can, in principle, be estimated in two ways: from trials like the 2 × 2 design above that experimentally manipulate expectancy, or from an RCT that only manipulates *A* (psilocybin/methylphenidate), by applying observational causal inference techniques.

To experimentally manipulate expectancy, one could deliver auxiliary sham treatments alongside the psychedelic/control. For example, one could randomize whether or not to deliver an inactive placebo like fish oil, paired with the *message* “this auxiliary drug has no effects on the psychedelic trip but may increase (decrease) the psychedelic’s anti-depressant qualities by enhancing (blocking) neurogenesis after the trip.” This has the advantage of altering expectancy, without the risk of acute harm to patients. By using a fake pill, it may also be a more efficacious expectancy manipulator. Since fish oil has putative neurogenesis effects (Wen et al., 2024), it may satisfy deception-related ethical concerns. Messages have also been proposed in vaccine RCTs to mitigate unblinding Stensrud et al. (2024); Obolski et al. (2024). If these or other existing strategies cannot perfectly experimentally set/manipulate expectancy levels, as some have suggested (Szigeti and Heifets, 2024), a contrast of average outcomes across arms for a fixed message will *not* correspond exactly to the CDE (see discussion of interpretations and statistical tools for such experiments in Appendix E.5). Nevertheless, even if expectancy levels cannot be perfectly set experimentally, assessing the joint impact of treatment and *practical* expectancy manipulations can help probe the impact of unmasking by testing the sensitivity of treatment effect estimates to interventions on expectancy.

Alternatively, the CDE can—under transparent assumptions—directly be estimated from even a standard trial (where only psilocybin/methylphenidate is randomized) using techniques from observational causal inference. Thus, we can estimate CDEs from trials that have already been conducted. A key assumption is that *E* is “as good as randomized” within levels of *X, A*, and *Z*. In the psilocybin example, it would likely be critical to include assessments of 1) anticipated pre-treatment hopes under both placebo and psilocybin in the vector *X*, and 2) immediate perceived effects of the treatment and (post-treatment) belief in the vector *Z*. For those that view expectancy effects as a key mechanistic component of psychedelics, one might also estimate the *treatment-fixed CDE of expectancy* (tfCDE), which quantifies the independent causal effect of expectancy within-arm. This is defined by the contrast 𝔼[*Y* (*a, e*) − *Y* (*a, e*^′^)] across two expectancy levels *e, e*^′^, for a fixed level of treatment *a*. In Appendix E, we state assumptions formally and discuss implications. Critically, the CDEs/tfCDEs cannot be estimated by stratifying analyses on expectancy/treatment levels or by including them as covariates in a regression model.

Importantly, the timing of expectancy measurements determines the interpretation of the CDE and the *E* → *Y* confounders that must be measured.

- **Pre-efficacy Effect Isolation** In some studies, the causal effect of pre-efficacy expectancies *E* (i.e., those occurring before onset of perceivable therapeutic benefits), can be isolated. This might be feasible if, for example, psilocybin’s anti-depressant effects were caused by neurogenesis processes that do not result in perceivable changes in depression until several days after treatment. Then expectancy levels measured a day after treatment could be conceptualized as pre-efficacy, and used to estimate a pre-efficacy CDE: the effect of the treatment under a fixed level of pre-efficacy expectancy changes. This would control for the effects of the “trip” on expectancies pre-efficacy, but not for post-efficacy expectancy changes from perceived therapeutic benefits.
- **Post-efficacy Effect Isolation** If therapeutic benefits are instead mediated through the trip, itself, then efficacy-expectancy feedback begins at trip onset (*t*_1_ = *t*^∗^) and one must therefore adjust for all common causes of post-efficacy expectancy and the outcome. In addition to pre-efficacy expectancy confounders, one would need to measure mediators of efficacy-expectancy feedback, like initial post-treatment measurements of *Y* (e.g., depression levels recorded after treatment but before the expectancy measurement). This formalizes how disentangling expectancy effects due to placebo effects versus perceptions of therapeutic benefits is more difficult post-efficacy.

In Appendix A.3, we discuss why we have centered our approaches for functional unmasking around CDEs/tfCDEs. For example, we describe why the well-known “Natural Direct Effect” is less relevant and unlikely to be identifiable in psychedelic studies (Robins and Greenland, 1992; Pearl, 2001). There are also related questions that correspond to estimands different from the CDE, such as *separable causal effects*. For example, suppose one sought to isolate distinct components of a psychedelic’s therapeutic effects attributable to the “the trip” versus non-psychoactive effects. This objective aligns with study designs that aim to “add the trip onto the control” (e.g., with active comparators) or “take the trip out of the drug” (e.g., coadministration of anesthesia (Lii et al., 2023b) or haloperidol (Vollenweider et al., 1998)). These designs implicitly rely on a conceptualization of the trip-related and non-trip-related effects as causally *separable* (Robins and Richardson, 2010; Robins et al., 2022; Stensrud et al., 2023, 2022), which would be justifiable if, for example, the components were generated by distinct neurobiological mechanisms.

### Statistical Mediation Analysis in Practice

Our proposed approach to expectancy-adjustment is based on modern causal inference methods that rely on fewer assumptions and have better statistical properties than classical regression-based approaches (see Appendix E.2 for a detailed explanation). This is critical as parametric approaches, like including confounders as covariates in a parametric model (e.g., linear/logistic regression), are only valid under strong, likely unrealistic assumptions. By using flexible machine learning methods as part of CDE estimation, one can capture complex interactions and non-nonlinearities without assumptions about the functional form of complex relationships between measurements of *Z* (e.g., trip “experience”), *X* (e.g., hype, set/setting), and *E*. We provide example code in Appendix F illustrating how to implement this mediation strategy. It also shows how our approach accurately recovers the true causal effect. Importantly, the method requires sufficient sample sizes, as theoretical guarantees are asymptotic.

### Sequentially Randomized Experiments

Sequential randomization of both expectancy manipulations and the treatment provides a strategy to probe the effect of treatment sequences that can answer questions related to dose, effect-durability, and expectancy, not possible with single timepoint designs. We illustrate a two timepoint design in Figure 5 for simplicity, although this approach can be applied with more timepoints. This generalizes parallel group dose-response designs. Results from such RCTs can be analyzed with *Marginal Structural Models* (MSMs), arguably the most popular modeling framework for longitudinal causal inference (Hernán et al., 2001; Robins et al., 2000; Robins, 1999) test the effects of specific treatment and message sequences (Robins et al., 2000; Robins, 1999). MSMs can also test the effects of treatment and expectancy sequences by applying the observational causal inference techniques discussed above, thereby echoing and providing a framework for the “call to action […] to thoughtfully integrate longitudinal assessments of expectations into future studies” (Colloca et al., 2023). The RCTs designs allow one to test how treatment and expectancy effects evolve across the study, potentially enabling one to reveal therapeutic benefits obscured by expectancy effects. For example, in Figure 6 we illustrate causal effects related to 1) how long therapeutic benefits of expectancy and/or the psychedelic last, 2) how the outcome changes depending on the number of treatments or expectancy manipulations, 3) the psychedelic’s effects on expectancy, and 4) the CDE. By randomizing assignment at multiple timepoints, participants have multiple opportunities for treatment and are not simply randomized to “one of two arms.” Thus the design may reduce participant pre-occupation with identifying their treatment arm, a common challenge in two-arm RCTs (Szigeti and Heifets, 2024). In Appendix E.6, we propose ways to quantify/test the causal effect of expectancy manipulations and treatment.

**Figure 5.**
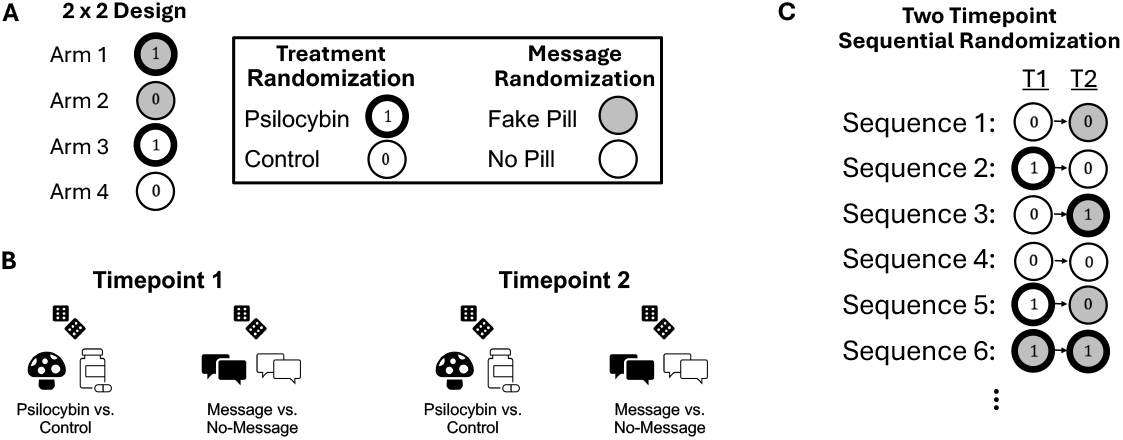
Sequentially Randomized Design. (A) Example 2×2 design at a single timepoint shows that for a single timepoint there are four possible arms/sequences from the treatment × message combination. At each timepoint, we indicate psilocybin with ➀ and control with 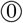. Similarly, we indicate expectancy manipulation message and no-message with gray and white, respectively. (B) In a two timepoint sequentially randomized experiment, we can separately randomize the treatment (psilocybin/control) and the message at each timepoint. These randomizations can be conducted marginally, or conditional on observed variables such as baseline covariates (e.g., age, sex). At a second timepoint, one could again randomize marginally, or conditional on the response of a participant (e.g., expectancy, depression levels) to previous treatments/messages. This could be used to, for example, increase the chances that psilocybin-treated participants exhibit comparable expectancy levels to control participants. (C) A two timepoint sequentially randomized experiment yields a collection of “arms”, each with different combinations/sequences (often called “regimes” or “policies”) of treatment and messages. “T1” and “T2” indicates timepoints one and two, respectively.

**Figure 6.**
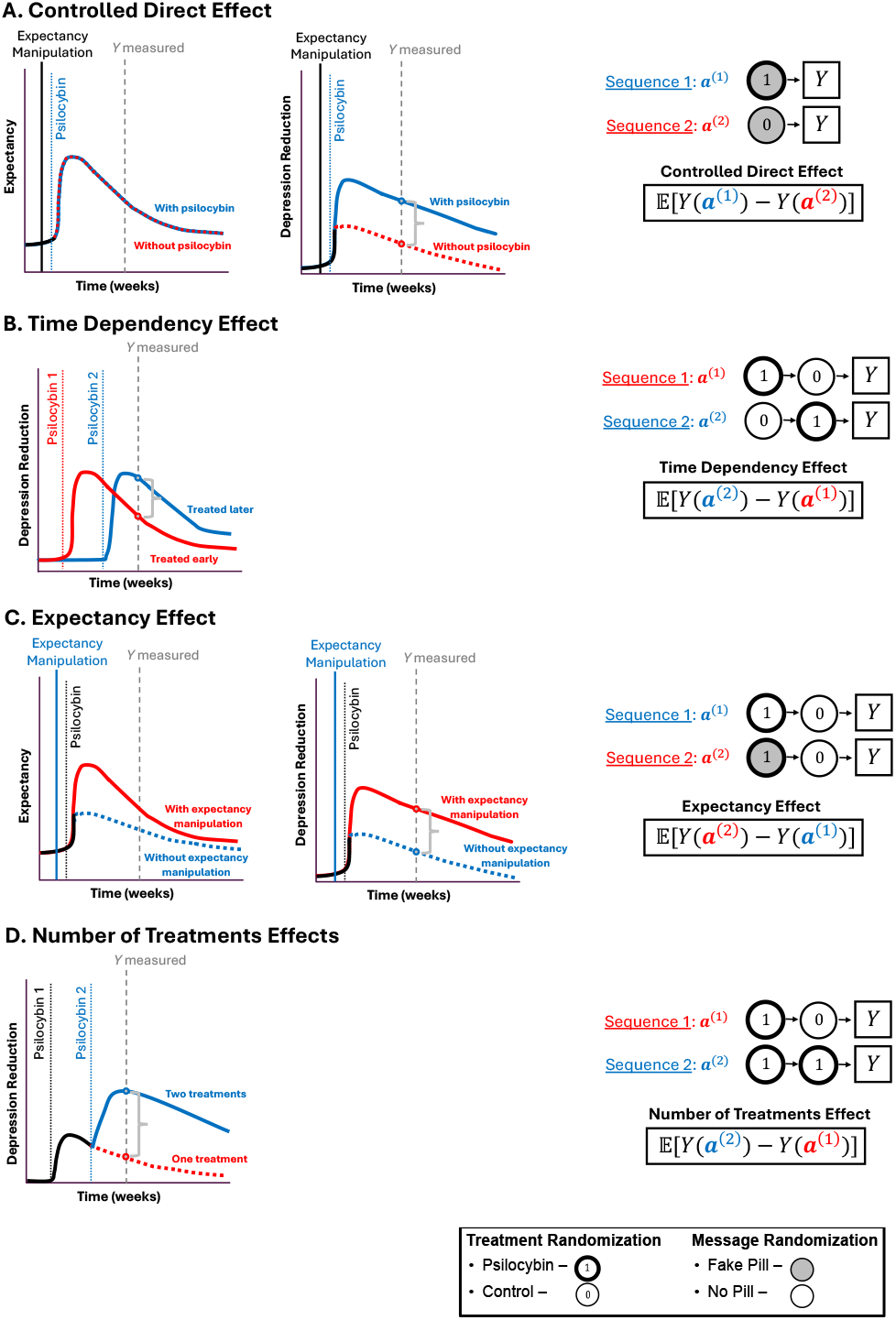
Example causal effects that can be tested in psychedelic RCTs. Each row contains (left) an illustration of potential causal effects on the outcome (depression) and expectancy (where relevant). On the right, we show treatment sequences/combinations (“regimes”/“policies”) of treatment/control and expectancy manipulation messages/no-messages, along side relevant causal effects (“estimands”) that can be estimated and tested with, for example, a Marginal Structural Model. We formalize these causal effects in Appendix E.6. The hypothetical depression and expectancy levels are shown in red/blue lines to differentiate different scenarios (e.g., with vs. without expectancy manipulation). Vertical lines indicate timepoints when expectancy manipulation messages and/or psilocybin treatment are administered, and *Y* is measured. We use a gray bracket measuring the distance between two dots to show the target causal effect to be estimated by the equation on the right. (A) Controlled Direct Effect in an idealized setting. The figures on the left illustrates the quantity targeted: a difference in depression levels between psilocybin/control arm participants (middle) in a world where the expectancy levels are identical in psilocybin and control arms (left). As indicated by the Treatment Sequences ***a***^(1)^, ***a***^(2)^, all participants in the sample are administered an expectancy manipulation message (gray fill), but includes both psilocybin (➀) and control arm 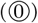 participants. (B) Time Dependency Effects compares the durability of a psilocybin treatment by comparing depression levels between subsets of participants who received treatment at either early or late timepoints. (C) Expectancy Effects captures the causal effect of expectancy manipulation vs. no-manipulation among a subset of participants administered psilocybin. (D) Number of Treatments effects quantifies the causal effect of repeated administrations of psilocybin (one vs. two). This strategy can be used to compare other contrasts (e.g., two vs. zero treatments).

### Causal Inference Complements Existing Functional Unmasking Strategies

We believe our modern causal approach can help investigators reason about statistical and design strategies to mitigate bias from unmasking. First, the causal framing highlights the goal of randomization which we believe deserves discussion. For example, Schenberg (2025) argues that RCTs do not deserve the “gold standard status” in part because variable “balance through random allocation […] is not actually granted” in any given sample. Our view is that randomization enables causal inference by ensuring the no unmeasured confounding assumption, not by inducing exact variable balance. While a lack of exact confounder balance between-arms can lead to small sample bias, this does not imply the treatment effect is confounded. Thus, as Schenberg (2025) rightly acknowledges, standard errors should capture uncertainty. Therefore, hypothesis tests of, for example, ATEs remain valid. While successful belief- and/or expectancy-blinding would lead to balance in post-treatment belief and/or expectancy values on average, balance between-arms is not enough to ensure that study results yield causal quantities that correspond to successful masking. This misconception has led to proposed remedies that we believe have drawbacks. For instance, Szigeti and Heifets (2024) propose a resampling strategy to generate adjusted samples with balanced expectancies across treatment arms. This illustrates the need for counterfactual reasoning: confounding occurs at the individual-, not sample-, level and thus reweighting observations to induce sample balance between arms will not resolve the problem. In Appendix E.4.1, we derive the causal quantity that the Szigeti and Heifets (2024) strategy targets and show how it can yield estimates that are so biased that they have the opposite sign of the true direct causal effect.

Second, the DAG suggests a framework for incorporating variables that may strongly influence psychedelic efficacy into analyses, such as, pre-treatment effect modifiers *X* (e.g., hype, set/setting), or post-treatment expectancy-outcome confounders (e.g., trip experience, post-treatment therapeutic alliance). For example, the FDA recently requested future MDMA applications (U.S. Food and Drug Administration, 2024) “incorporate an assessment” of hype. But how? The DAG illustrates that hype is a potential confounder of the *E* → *Y* relationship, and thus should be included in the pre-treatment covariate vector *X*. However, while hype may modify the *A* → *Y* effect, it is *not* a treatment-outcome confounder. Some have suggested running RCTs among low hype participants (Nayak and Zahid, 2025), or using hype as a covariate in a regression model (Aday et al., 2022) in settings prone to functional unmasking. We agree that these strategies will provide valuable insight but we believe that they will not, in general, fully resolve the challenges of unmasking. Such strategies would instead target a *conditional* average treatment effect (CATE): the ATE in a subpopulation that has a specific hype level. Decision-making from RCT results is rarely based on this type of CATE because they may not generalize to other populations. For example, perhaps psychedelics are beneficial among those with positive/neutral pre-treatment attitudes but damaging to those with negative attitudes because they have more bad trips. We discuss this further in Appendix B.

Third, our work clarifies why previous statistical proposals in psychedelic RCTs may not alone resolve challenges from unmasking. For example, Muthukumaraswamy et al. (2021) propose, what we believe, is a *principal stratum* strategy (see expression 14 in their publication). This paper has many virtues, but we believe the principal stratum causal quantity has considerable drawbacks (see, e.g., Stensrud et al. (2023)). Their approach yields estimates that are interpreted as the causal effect of treatment among participants who *would have* the same level of belief in both arms (i.e., the people who *would be* successfully belief-blinded). The downside is that this subpopulation may not exist, and even if it does, it may be highly unrepresentative of the general population. For example, the principal stratum estimate may reflect the psychedelic’s causal effect among people for whom treatment had little therapeutic benefit, thereby explaining why they meet the *would-be successfully belief-blinded* criteria. Moreover, the proposal of Muthukumaraswamy et al. (2021) relies on highly parametric assumptions that may be unrealistic in this setting. We discuss other proposals in Appendix E.3.

Fourth, our approach reveals how RCT design strategies can be improved by explicitly stating causal estimands, corresponding to hypothetical target trials. For example, dose-response psychedelic designs (Goodwin et al., 2022) and/or use of low-dose psychedelics as controls may not alone resolve the expectancy-mediated challenges of functional unmasking because expectancy levels may exhibit a dose-response relationship with the psychedelic dose as we illustrate in Figures 2F-G. That is, the ATE may grow with dose only because expectancy-mediated effects increase with dose. However, testing whether a CDE grows with dose is likely a powerful way to mitigate challenges from unmasking. Similarly, causal theory also explains why “deep blinding” (Matvey et al., 2025; Barrett et al., 2018) may not fully resolve functional unmasking. Deep blinding (Matvey et al., 2025; Barrett et al., 2018) refers to strategies whereby control participants receive an active comparator (e.g., methylphenidate), treatment participants get active treatment (e.g., psilocybin), and all participants are led to believe they could receive any drug from a long list. While deep blinding may lead to belief-blinding, it does not ensure expectancy-blinding (as emphasized in the DMT-salvia example). Thus, while active comparators are currently selected to achieve belief-blinding, the DAG suggests that they should be chosen to induce comparable expectancy levels as the treatment (expectancy-blinding). We formalize this in Appendix E.7. Counterfactual theory also provides technical rationale for deep blinding: it ensures that the *positivity* /overlap causal assumption is met by encouraging sufficient variability in expectancy levels across, for example, treatment arms (discussed in Appendix E). Thus, these designs are remedies for unmasking, when integrated with the formal causal approach.

By pairing an estimand-first strategy with the estimation methods discussed, investigators can rigorously define and estimate causal quantities extending far beyond comparisons of average outcomes across treatment arms. This aligns with the idea of target trials. Indeed, the ICH E9 guideline on statistical principles for RCTs (Kahan et al., 2024) encourages trialists to specify the scientific question in terms of causal estimands and assumptions prior to study design. This contrasts with a more indirect approach that starts with design considerations (e.g., by asking “what is a good control condition for that treatment?” (Colloca and Fava, 2024)). Together, we hope our work provides a basis for strategies that mitigate the unwanted effects of functional unmasking and improve the rigor of RCTs for psychedelic and mental health treatments, more broadly.

## Data Availability

No data were collected or analyzed in this work. Code for our example analysis guide and for reproduction of simulations/figures are available at GitHub: https://github.com/gloewing/RCTs_unmasking

## Acknowledgments

This research was supported in part by the Intramural Research Program of the National Institutes of Health (NIH) (ZIC MH002968). The contributions of the NIH author(s) were made as part of their official duties as NIH federal employees, are in compliance with agency policy requirements, and are considered Works of the United States Government. However, the findings and conclusions presented in this paper are those of the authors and do not necessarily reflect the views of the NIH or the U.S. Department of Health and Human Services. We thank Dr. Francisco Pereira, the NIMH Machine Learning Core, and the members of the Center for Psychedelic and Consciousness Research for their helpful suggestions and feedback on manuscript drafts. Mats J. Stensrud was supported by the Swiss National Science Foundation (Starting Grant 211550).

## Appendices

### A Prior conceptualizations and methodological work

#### A.1 Comparison between belief/expectancy definitions in our proposal and prior work

Our expectancy definition is similar to a previously proposed “outcome expectancy” definition (Tambling, 2012). The belief/expectancy distinction is close to those proposed in (Butler et al., 2022; Szigeti and Heifets, 2024), but differs from the Evidence Based Medicine literature (e.g. see Chapter 6.3 of Howick (2011)).

#### A.2 Connection to Past Statistical Methods Work on Unmasking

Our approach builds on the work of Liu et al. (2016); Jamshidian et al. (2014); Hubbard et al. (2012); Zhang et al. (2013) that propose targeting controlled direct effects using flexible mediation analysis techniques, and mediation analyses in randomized trials (Lynch et al., 2008). Gabriel et al. (2025) conceptualize the problem in terms of natural direct effects.

In contrast to all the above works, we propose to use expectancy as the key mediator rather than belief, and we identify time windows across which confounders may vary. Moreover, our conceptualization suggests feedback mechanisms that provide potential explanations for seemingly unintuitive null results in psychiatry research and suggests more precisely what confounders must be measured to isolate treatment effects that are not mediated through expectancies. We consider statistical strategies proposed in the psychedelic literature for functional unmasking in Appendix E.3. Other reviews (Ohlsson and Kendler, 2020) have considered the use of observational causal inference methods in psychiatry research.

#### A.3 Natural Direct Effects

We do not pursue the natural direct effect (NDE) as a target for mediation effects, mostly due to conceptual problems with these estimands that limit their practical relevance. First, these effects are so-called “cross-world”: they are defined with respect to interventions that cannot be implemented in real-life, even in principle (Richardson and Robins, 2013). Second, and relatedly, identification of these effects require assumptions that are neither testable nor experimentally enforceable, even in principle. Importantly, controlled and separable effects do not suffer from these drawbacks. Recent work in causal inference has suggested alternatives to natural effects, often called randomized intervention analogues (VanderWeele et al., 2014). However, the stochastic interventions invoked in the definition of these effects also have a dubious practical relevance, see e.g., Miles (2023); Díaz (2024); Hernan and Robins (2024). More broadly, attempts to justify the relevance of natural effects have often been based on stories corresponding to different estimands (Robins and Richardson, 2010; Stensrud et al., 2023), now often called separable effects.

Separable effects may be useful in psychedelic research as a tool to combat functional unmasking, or to aid in the development of new compounds. For example, to motivate these effects, consider a drug developer who aims to create a new therapeutic compound that has no (“psychoactive”) effect on expectancy, but has a (“therapeutic”) effect on the outcome that is direct, analogous to the direct effect of psilocybin. A separable direct effect would simply compare this hypothetical drug to current psilocybin. Because this drug does not exist, but might be developed if the current data suggest that it could have promising effects, it is of interest to identify and estimate this effect from existing data, where only psilocybin and placebo are available. Indeed, if, based on the analysis of existing data, we expected this new drug to have a large effect, it would also signal that there is a “therapeutic” component in psilocybin that is active. Thus, separable effects may be of interest purely from a explanatory/mechanistic perspective, even outside of any pursuit of more targeted future interventions.

Separable effects can be estimated from RCT data using assumptions that differ from those needed for controlled direct effects. Like for the controlled direct effect, we need to adjust for common causes of exposure and outcome. Furthermore, we need to carefully describe how any post-treatment mediator–outcome confounders are affected by psilocybin, and clarify whether we want the new treatment to affect these post-treatment mediators or not.

To explain the assumption intuitively, suppose hypothetically that psilocybin’s effects on brain-derived neurotrophic factor (BDNF) were entirely responsible for its therapeutic benefits, and its expectancy effects were entirely caused by psilocybin’s effects on the 5HT2A receptor. In this scenario, one could block the 5HT2A receptor and the resulting expectancy effects, without any alteration of psilocybin’s therapeutic properties. Similarly, one could block BDNF, and the resulting therapeutic effects, without any alteration of psilocybin’s expectancy effects. This is, notably, distinct from the reasoning behind a controlled direct effect, that allows for these processes/components to be intertwined, but targets a treatment effect at a fixed expectancy level. Finally, we note that, as alluded to above, identification of separable effects also requires careful consideration of how post-treatment mediator-outcome confounders are affected by the two mechanistic elements of the treatment—see Stensrud et al. (2023) for details.

### B Pre-treatment Expectancy

The DAG underscores how to use pre-treatment expectancy in analyses: it can be an effect modifier of the *A* → *Y* pathway, but it is not a treatment-outcome confounder because randomization ensures there are no causes of *A* and thus no common causes (confounders) of both *A* and *Y*. Similarly, pre-treatment expectancy is unlikely to be the *only* common cause of post-treatment expectancy, *E*, and depression levels, *Y*. We contend that conditioning on baseline expectancy (Szigeti et al., 2024b) is valid and can answer interesting questions but does not on its own account for functional unmasking without additional and likely unrealistic assumptions, which we detail below. While baseline expectancy is often predictive of treatment effects (Kaertner et al., 2021), conducting stratified analyses, or including it as a term in a regression, implicitly targets a *conditional average treatment effect* (CATE): the treatment effect in a sub-population defined according to baseline covariates (e.g., the treatment effect among participants who report low baseline expectancy levels). Treatment effect estimates in an RCT conducted among, for example, a population with low pre-treatment hype, would also yield a CATE (i.e., the treatment effect in a population with low pre-treatment hype). Decision-making from RCT results is rarely based on this type of CATE because they may not generalize to other populations.

If one assumed that unmasking was not a threat within a specific covariate stratum (e.g., participants with low baseline expectancy), then the CATE may quantify the direct effects of the treatment on the outcome in that stratum (i.e., effects not mediated through *E*). The degree to which such effects would generalize to other subpopulations, however, is entirely unclear. Alternatively, it may be intuitive to conceptualize post-treatment expectancy as largely determined by pre-treatment hype. In other words, one might argue that the trip *A*—and its immediate effects on belief about treatment received, *B*—”unlocks” one’s pre-treatment expectancies: if one believes they *would* benefit from the treatment before the trial, then unmasking may result in a participant expecting *they will* benefit from the treatment they just received. Thus, under this model, unmasking “unlocks”/”unleashes” their pre-treatment hype. If indeed this were the entire mechanism by which post-treatment expectancies *E* were determined, a CATE would implicitly target a conditional controlled direct effect (cCDE), which is a valid mediation-style estimand, but again defined within a particular covariate stratum. In reality, however, we believe pre-treatment hype may contribute but is *not* the sole common cause of post-treatment expectancy, *E*, and the outcome, *Y*. For example, participants with low pre-treatment expectancy may experience a mystical experience and result in high post-treatment expectancy. Similarly, some participants with high pre-treatment hype may experience a weak or scary trip, ultimately resulting in low post-treatment expectancies. Thus, it is very unlikely that post-treatment *E* is solely caused by pre-treatment hype and belief *B* because *A* is also a common cause of post-treatment *E* and the outcome (e.g., potentially through intermediate variables *Z*). Thus for multiple reasons, basing decisions on a CATE estimate will *not* resolve the challenges of functional unmasking. We show in the main text how to use baseline expectancy to adjust for expectancy effects in a formal manner.

### C Blinding Types

Here we describe types of blinding in terms of causal pathways.

### D Longitudinal Description

We begin with a longitudinal version of the DAG presented in main text 1A and a high level description of efficacy-expectancy feedback. We then describe these in greater depth below. The DAG shown in Figure 7 is a two timepoint extension that, although simplified, allows for visualization of key elements of the efficacy-expectancy feedback process. The variables *M*_1_ and *M*_2_ are mediators of efficacy-expectancy feedback. For example, *M*_2_ would be unprotected sexual behavior at timepoint *t* = 2 in the STI vaccine example in Figure 3. The test results, given each week during the study, might “scare” a participant into lowering their risky sexual behavior if they test positive. This in turn influences future test results, expectancies and beliefs. Similarly, in the Aspirin example in Figure 3, *M*_1_ might be the perceived headache level at timepoint *t* = 1. Perceived headache levels in the middle of the study then influence beliefs about the treatment received (*B*_1_) and expectancies about how the headache will improve (*E*_1_). These expectancies and beliefs in turn influence future headache level perceptions (e.g., *M*_2_).

**Figure 7.**
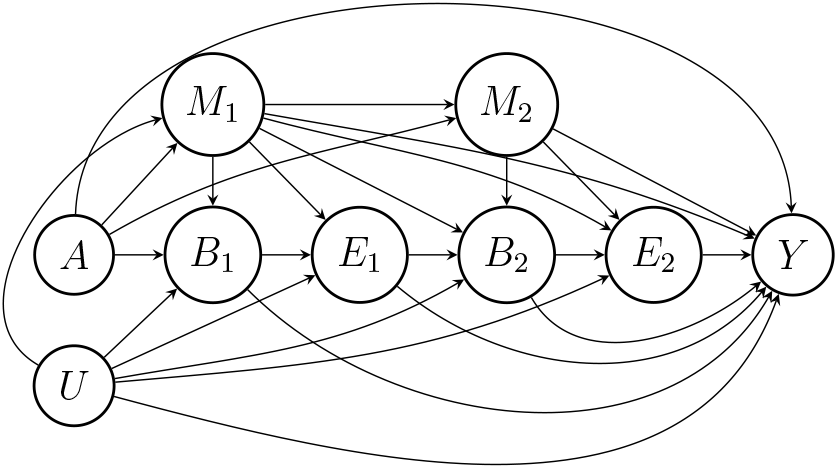
Simplified two timepoint DAG illustrating efficacy-expectancy feedback. For visual clarity, we represent a DAG that is within levels of baseline variables *X*. The full two timepoint DAG would have *X* point to all other variables except *A*, which we assume is randomized marginally.

#### D.1 Efficacy–Expectancy Feedback

In this appendix section, we present a more detailed version of the longitudinal conceptualization of RCTs presented in the main text. First we start with the sequence of hypothetical studies (illustrated in Figure 8) to differentiate between 1) unmasking from perceivable sideeffects versus from therapeutic benefits, and 2) expectancies before/after perceived benefits, or “efficacy.” We conceptualize intermediate variables on the *A* → *Y* pathway as stochastic *processes*, indexed by time, *t*. Let 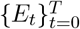 and 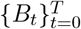 denote potentially latent/unobserved values of expectancy and belief be stochastic processes. Henceforth, variables indexed at *t* = 0 (e.g., *B*_0_, *E*_0_) are conceptualized as those measurable just prior to initialization of the trial (“baseline”), and *t* ∈ {1, …, *T* } denotes timepoints after treatment administration. We define 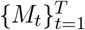 to be the values of the (potentially latent) stochastic process of “non-nuisance” mediators of treatment efficacy—these are made more concrete in each of the examples below. The pre-/post-expectancy efficacy distinction extends the “malicious” and “benign” unmasking concepts (Howick, 2011) and illustrates that post-efficacy expectancy (which would be categorized as “benign” in the taxonomy of (Howick, 2011)) can be a nuisance, and thus can make the classic RCT efficacy summary, the Average Treatment Effect (ATE), a less desirable statistical target in some designs. The ATE in these studies is 𝔼[*Y* (*a* = 1) − *Y* (*a* = 0)]: which can often be estimated as the difference in average outcome values across arms. We first give a concrete formalization to temporal feedback between expectancy and treatment efficacy, then illustrate this concept through describing RCTs with and without this potential nuisance pathway.

**Figure 8.**
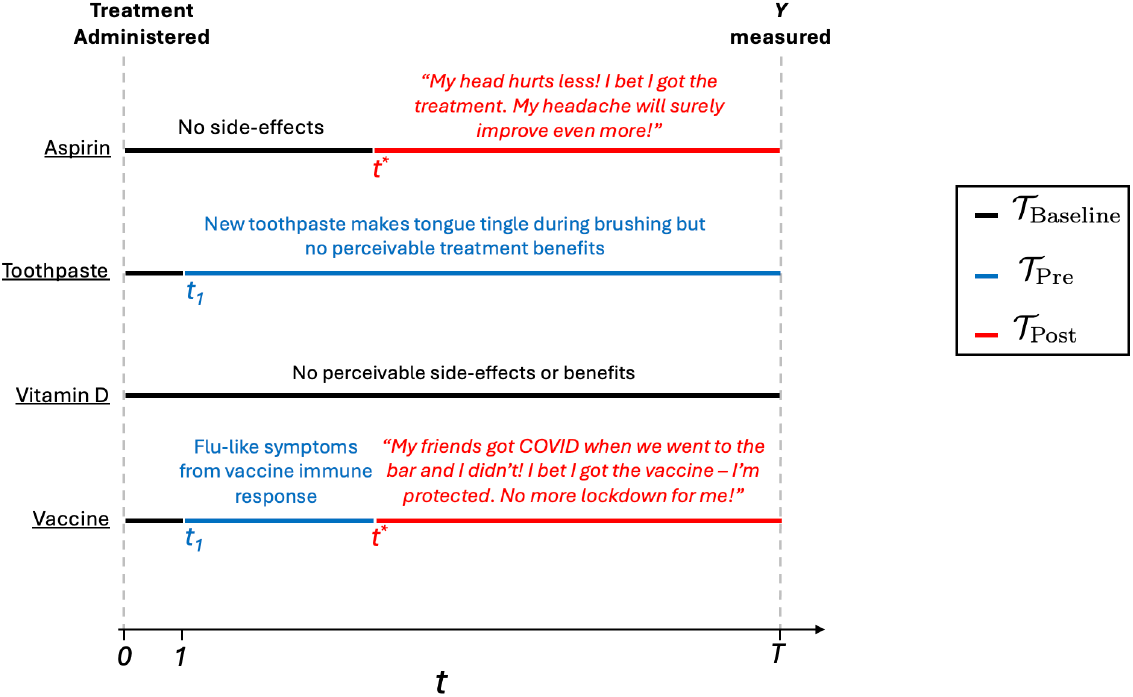
Example RCTs with and without side-effects and/or efficacy-expectancy feedback. *Aspirin* has no perceivable effects until *t*^∗^, the timepoint participants begin to feel improvements in their headache. Thus, 𝒯 _Baseline_ ={ 0, 1, …, *t*^∗^ − 1 }and 𝒯 _Pre_ =. ∅ The perceptible treatment effect starting at *t*^∗^ induces efficacy-expectancy feedback and thus 𝒯 _Post_ ={ *t*^∗^, …, *T* }. The *New Toothpaste* has side-effects (e.g. the tongue “tingles”) whenever it is “administered” (starting at *t*_1_ = 1) that the comparator toothpaste does not and so 𝒯 _Baseline_ = {0 } and 𝒯 _Pre_ ={ *t*_1_, …, *T* }. However, treatment benefits to gum health are not perceptible during the study and so there is no efficacy-expectancy feedback: 𝒯 _Post_ = ∅. The *Vitamin D Supplement* has no perceivable effects during the study and thus 𝒯 _Baseline_ = {0, 1, …, *T* } and 𝒯 _Pre_ = 𝒯 _Post_ =. ∅ The *COVID-19 Vaccine* induces flu-like symptoms the day after administration because of the acute immune response starting at *t*_1_ = 1: 𝒯 _Baseline_ = {0 } and 𝒯 _Pre_ = {*t*_1_, …, *t*^∗^ − 1 }. About six weeks after administration, treatment arm (*A* = 1) participants notice that they did not develop COVID symptoms when exposed, but their unvaccinated friends got sick. As a result of their perceived protection, participants begin taking more risks (e.g. going to restaurants more often). This induced efficacy-expectancy feedback means the set 𝒯 _Post_ = {*t*^∗^, …, *T* }. Table 2 describes these sets in terms of a 2 × 2 table of side-effect timing vs. efficacy-expectancy feedback.

##### Pre-vs. Post-Efficacy Expectancy

Side-effects may influence participant expectancy before the treatment has begun to exert its therapeutic effect. This is arguably one of the most common concerns surrounding “functional unmasking.” For that reason, we introduce notation to distinguish between time-points before and after *t*^∗^, the first timepoint that expectancy may be affected by these intermediate therapeutic effects, *M*_*t*_. We take, without loss of generality, *t* = 0 to be the baseline study timepoint when there are no paths from *A* to any other variables, measured or unmeasured (e.g. at study intake). We also define *t*_1_ ∈ {1, …, *T* } to be the first timepoint when there exists a path *A* → *E*_1_, or *T*, whichever occurs first. The 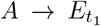 path may be mediated through the process 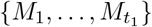 if, for example, there are no side effects, and expectancies differ between treatment arms at *t*^∗^ only because of efficacy-expectancy feedback (e.g. as in the Aspirin example above). On the other hand, if there are side-effects that occur prior to the onset of any therapeutic effects, then the 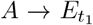 path would not be mediated through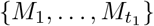. Formally, we define *t*^∗^ as the first timepoint where there exists a path 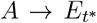 mediated through 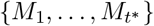, or *t*^∗^, whichever occurs first. We partition the post-treatment study timepoints 𝒯= {1, …, *T* } into three subsets: (i) the (potentially empty) set of timepoints prior to any effects on expectancy, 𝒯 _baseline_ = {1, …, *t*_1_ − 1}, (ii) the (potentially empty) set of timepoints from *t*_1_ up to *t*^∗^, 𝒯 _Pre_ = { *t*_1_, …, *t*^∗^ 1 } and (iii) the timepoints from *t*^∗^ onwards, 𝒯 _Post_ = { *t*^∗^, …, *T* }. We provide examples of these sets in Figure 3 and Table 2 for common RCT settings.

##### Aspirin RCT

Imagine we run a double blind RCT to study the efficacy of aspirin, relative to an inactive control, to treat headaches. The study achieves *complete blinding* prior to the onset of any treatment effects as *A* has no perceptible effects except for its impact on intermediate perceived headache levels, 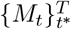. One hour after administration, *A* = 1 arm participants perceive a headache reduction resulting in a “snow-balling” effect: the therapeutic benefit leads to greater expectancy (i.e. *A* → *M*_*t*_′ → *E*_*t*_′). The patient thinks “my headache is improving, I bet I got the active treatment, and my headache will improve even more.” This greater expectancy further improves the perceived efficacy of the treatment, which again feeds back leading to even greater expectancy levels (i.e., *A* → *M*_*t*_′ → *E*_*t*_′ → *M*_*t*_′_+1_→ *E*_*t*_′_+1_→ …). We omit intermediate values of *B*_*t*_ for notational ease, with the understanding that mediation through *E*_*t*_ determines the identifiability of isolated effect.

The above example describes *efficacy–expectancy feedback*, which is illustrated in DAG 7. ATE estimates from RCTs which are classically understood as successfully double blinded may include effects mediated through efficacy–expectancy feedback: the effects of *A* on intermediate *M*_*t*_ are large enough to be perceivable to study participants prior to measurement of *Y*. Efficacy–expectancy feedback can be cast as a special case of treatment-confounder feedback from the sequentially randomized experients causal inference literature (Hernan and Robins, 2024). We use the graphical shorthand, *M*_*t*_ ⇋ *E*_*t*_ to denote this feedback process, with the understanding that the elaborated graphical model is acyclic.

##### Vaccine RCT

In Figure 4 we illustrate a case where efficacy-expectancy feedback is a nuisance. Imagine a study for a vaccine of for the sexually-transmitted infection Chlamydia among freshman college students on a campus with an outbreak. All freshman students are randomized to receive either the vaccine (*A* = 1) or a control (*A* = 0) shot before the first semester. We assume there are no side effects and that successful belief-blinding is achieved throughout. Participant infection status is tested weekly, and infected participants are given an antibiotic to treat the infection. For simplicity, imagine the antibiotic works instantaneously, and does not prevent future infections. At the end of the study, the average number of infections each participant experienced is compared across arms. Figure 4 illustrates how, even without vaccine side effects, efficacy-expectancy feedback can obscure strong treatment effects: testing positive for Chlamydia scares students into reducing their tendency to have unprotected sex. If the vaccine is effective, *A* = 1 students become infected with a lower probability for each exposure, and so over time, they exhibit riskier behavior than controls on average (i.e., vaccine participants are scared off from unprotected sex less than controls). Defining *M*_*t*_ as an indicator of unprotected sex at time *t*, we encounter the efficacy-expectancy feedback as above: *A* → *M*_*t*_′ → *E*_*t*_′ → *M*_*t*_′_+1_ → *E*_*t*_′_+1_ →… We formalize this in Appendix D.2 with a simulation study that mathematically expresses this feedback mechanism. A scientist might want to know what a *risk-controlled* effect of the vaccine is: does the vaccine reduce infection if treatment and controls have identical risky sexual behavior across treatment arms.

The above two examples illustrate that the ATE may not be the only statistical target; a controlled-direct effect may be preferable due to isolation of a nuisance pathway. This sheds light on other psychiatry RCT results. For example, perhaps SSRIs show only modest treatment effects because depressed participants experience the benefits of SSRIs and reduce behaviors they used to rely on to regulate their mood (e.g. exercise, meditation, therapy) because they feel the medication is now enough. Thus the medication helps reduce their depression, but over time, the decrease in these other behaviors increases their depression, leading to a diluted treatment effect. These examples illustrate that “pre-efficacy/post-efficacy” may be a better dichotomy than benign/malicious unmasking, defined as unmasking from perceived efficacy, or side-effects, respectively (Howick, 2011). Unmasking due to perceived treatment efficacy is considered “benign”, but as shown above, it can result in effect cancellation in a way that obscures the scientific question. In some policy cases, the ATE may represent “realworld impact.” In other cases, these feedback mechanisms may prevent assessment of scientific questions like “how does the pharmacology of psilocybin lead to a reduction in depression?”, which is important if one is developing an analogue chemical with therapeutic effects but no hallucinogenic properties. If psilocybin’s anti-depressant effects are largely mediated through expectancy, then such designer drug efforts are futile. Together these examples show that mediation of treatment effects through unwanted nuisance pathways may exist even if participants are belief-blinded, and clarifies that post-efficacy unmasking can be a nuisance.

#### D.2 Vaccine Simulation

Here, we describe a simple data generating scenario corresponding to the vaccine example in main text Figure 4, which is copied below here for convenience.

We will suppose that the relevant data comprise baseline treatment status *A* along with longitudinal measurements 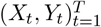 for each individual, where *X*_*t*_ is the risk factor status at timepoint *t* and *Y*_*t*_ is an indicator for infection at the *t*-th timepoint. In particular, we consider the following generative model:

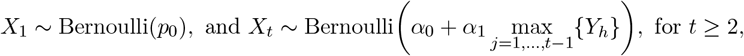

and

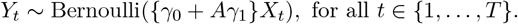

That is, we consider the following parameter values:

- *p*_0_ = ℙ(*X*_1_ = 1) = 1*/*2
- *γ*_0_ = ℙ(*Y*_*t*_ = 1 | *X*_*t*_ = 1, *A* = 0) = 3*/*4
- *γ*_1_ = ℙ(*Y*_*t*_ = 1 | *X*_*t*_ = 1, *A* = 1) − ℙ(*Y*_*t*_ = 1 | *X*_*t*_ = 1, *A* = 0) = −3*/*8
- *α*_0_ = ℙ[*X*_*t*_ = 1 | *Y*_1_ = · · · = *Y*_*t*−1_ = 0] = 1*/*2
- *α*_1_ = ℙ[*X*_*t*_ = 1 | ∃1 ≤ *j* ≤ *t* − 1: *Y*_*j*_ = 1] − ℙ[*X*_*t*_ = 1 | *Y*_1_ = · · · = *Y*_*t*−1_ = 0] = −2*/*5
  - With efficacy-expectancy feedback (Figure 9 [Left]): *α*_1_ = 1*/*2
  - Without efficacy-expectancy feedback (Figure 9 [Right]): *α*_1_ = 0

**Figure 9.**
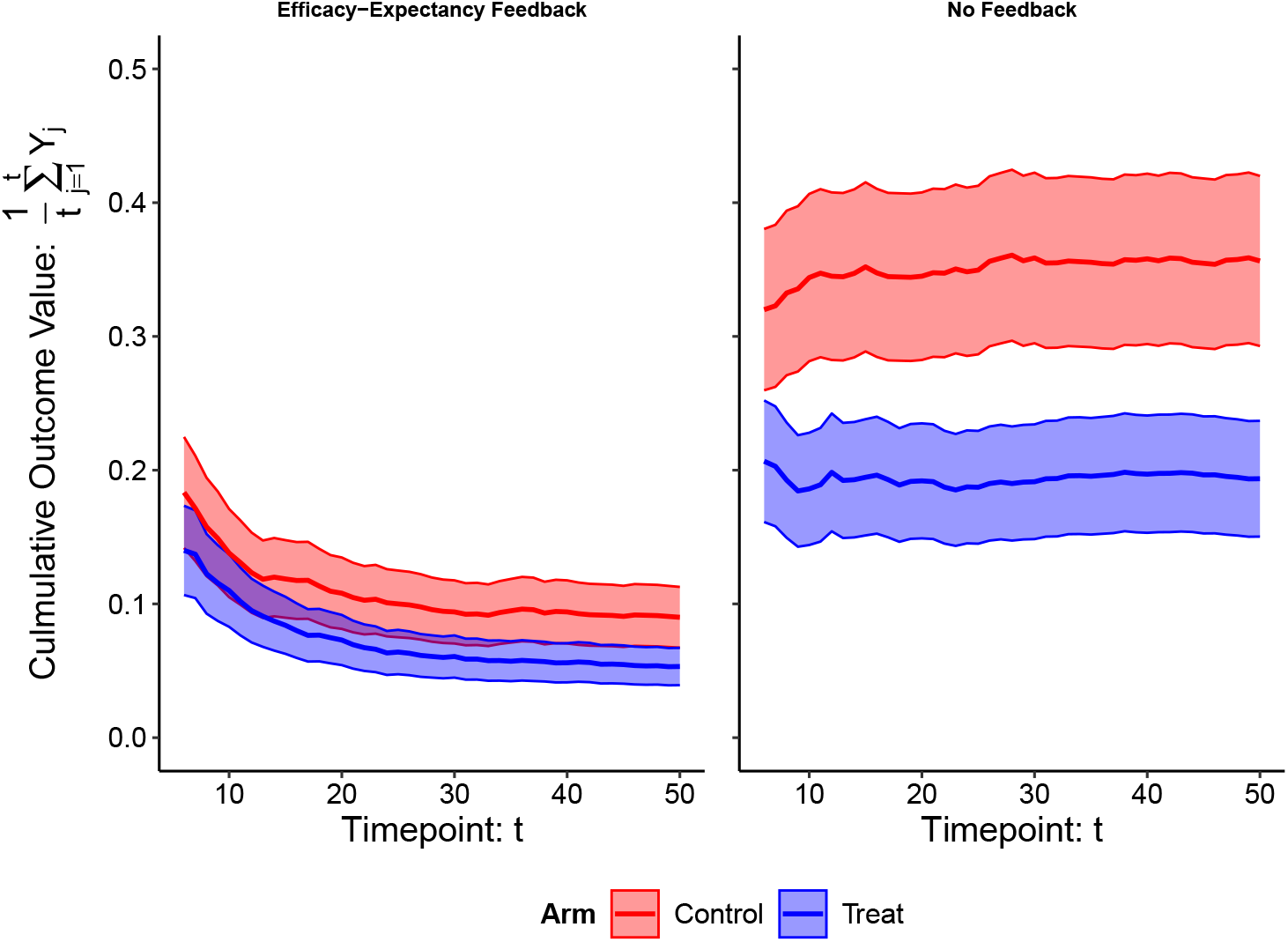
Efficacy–Expectancy Feedback Example. Vaccine RCT simulation example. [Left] Simulation results *with* efficacy-expectancy feedback: participants reduce their sexual risk behavior if they become infected. [Right] Simulation Results from an identical simulation but *without* efficacy-expectancy feedback: participant infection throughout the study does *not* alter their risk behavior. When participants have the same risk behavior in both arms: the vaccine reduces the risk of infection for each exposure by 50%.

If a participant has unprotected sex at timepoint *t*, then their chances of becoming infected are *γ*_0_ = 3*/*4 in the control arm and *γ*_0_ +*γ*_1_ = 3*/*4 − 3*/*8 = 3*/*8 in the active vaccine (treatment) arm. Thus the vaccine is imagined as highly protective in this simulation: it cuts the risk of infection (upon exposure to the STI) at any timepoint in half. This example is not to imply any such vaccine for this disease exists, but rather to illustrate that even if a treatment is highly effective, efficacy-expectancy feedback can obscure the effectiveness. The mean of *X*_*t*_ is a function of the 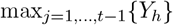. This describes in this scenario how participants reduce their risk if they have been infected at any point in the study prior to the current timepoint *t* (i.e., they get partially “scared” off from having unprotected sex if they get infected at least once). Participants who have never been infected have a probability of *α*_0_ = 1*/*2 of having unprotected sex. On the left we show the case where efficacy-expectancy feedback exists: if participants become infected at any point, their chances of having unprotected sex in their future fall to *α*_0_ + *α*_1_ = 1*/*2 − 2*/*5 = 1*/*10. On the right, we show the same simulation in a hypothetical study where there is no efficacy-expectancy feedback: regardless of whether a participant has been infected at any past study timepoints, their probability of having unprotected sex at future study timepoints is the same *α* = 1*/*2. Together this figure illustrates how efficacy-expectancy feedback can obscure treatment effects.

### E Formalizing Mediation Effects

To estimate the controlled direct effect (CDE), we require three key assumptions alluded to in Section: (i) *consistency*, (ii) *positivity*, and (iii) *no unmeasured confounding*. We will consider these in the context of a standard randomized controlled study where only *A* is randomized, though these assumptions remain relevant in other study designs. The three causal assumptions for mediation are formalized as follows:

**Assumption 1** (Consistency). *Y* (*a, e*) = *Y whenever A* = *a and E* = *e*.

**Assumption 2** (Positivity). *For some ϵ >* 0, *the following holds for each* (*a, e*) *pair:*

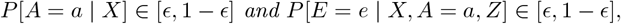

*with probability 1*.

**Assumption 3** (No unmeasured confounding). *For each* (*a, e*) *pair*,

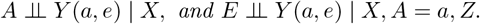

First, consistency (Assumption 1) essentially requires that the intervention on *A* and *E* is well-defined, and that there is no interference between subjects, i.e., that treatment of one study participant does not affect the outcome of another participant. A bit more formally, consistency states that whether one *intervenes* to set *A* and *E* to specific values, or these values *happen to occur* in the real world, the value of the resulting outcome should be the same. For treatment *A*, this will typically be satisfied in a well-designed study, so long as there is no interference between participants—this might fail if study participants are connected via social networks and are influencing each others’ behaviors in response to their treatment. For expectancy *E*, while considerations of interference are similar, the notion of a hypothetical well-defined intervention is more subtle. To be concrete, suppose that the gold-standard measurement of post-treatment expectancy *E* (e.g., the Credibility and Expectancy Scale (Devilly and Borkovec, 2000)) is captured through responses to an instrument/questionnaire. The notion of intervening on *E* and setting it to the value *e* can be interpreted as a hypothetical intervention where a trusted physician converses with the subject after their treatment experience and convinces them to achieve specific targeted answers *e* to each component of the questionnaire. Moreover, the measurement of *E* through, say, a questionnaire, should be sufficiently rich such that the same value of *e*—regardless of how it is achieved—always represents the same effect on outcomes. This last criterion is, perhaps, less critical, as a valid causal interpretation is still possible even when it fails (VanderWeele and Hernan, 2013).

Secondly, the no unmeasured confounding assumption (Assumption 3) requires us to measure all common causes of treatment and the outcome, as well as those of expectancy and the outcome, *Y*. When treatment is randomized (either marginally or within levels of measured covariates), the former requirement will hold by design. Moreover, when *only* treatment is randomized, the latter requirement for measuring all confounders of the expectancy-outcome relationship is a substantive, untestable assumption. More concretely, we need to measure pre-exposure variables *X* and post-exposure variables *Z* that affect both *E* and *Y*, such that within levels of *X, A*, and *Z*, expectancy *E* is “as good as randomized”. In the psilocybin example, it would likely be critical to include in *X* a baseline assessment of anticipated expectancies under both placebo and psilocybin, and in *Z* one’s assessment of the immediate perceived effects of the treatment as well as a measure post-treatment belief, especially if the latter may influence outcomes directly and not through *E*.

Lastly, and critically, the positivity assumption (Assumption 2) dictates that within levels of these confounders, there must be sufficient variation in treatment and expectancy levels. When treatment is randomized this will again typically hold for treatment by design (e.g., if all treatment levels occur with equal probability). For expectancy, however, we need to ensure that *X, A* and *Z* do not rule out any values of *E*. For example, if belief is a confounder, then we require that belief does not systematically determine expectancy. Similarly, we must ensure that within each treatment arm, there is sufficient variability in expectancy levels. This provides a clear counterfactual rationale for why “deep blinding” strategies (Matvey et al., 2025; Barrett et al., 2018) are likely important: they can help ensure that within levels of belief and treatment arm, there is sufficient variability in expectancy levels. This also has implications for the conceptualization and measurement of other post-treatment variables *Z*. For example, one may want to avoid questions such as “Did you have a good trip?”, which may overlap significantly with *E*, and instead opt for wordings similar to “Did you perceive noticeable immediate effects of the pill?” or “Did you hallucinate?”.

#### E.1 Effect Isolation

##### Post-efficacy Effect Isolation

We further discuss pre-efficacy effect isolation assumptions. Because of the positivity assumption outlined above, it is critical that all variables (*X, Z*) be distinct enough from *E* so as to ensure sufficient variability in *E* levels within all strata of these variables. For example, if one used large language model-based numeric representations of unstructured interviews about the acute trip, analysts would have to ensure these summaries do not accidentally capture qualitative properties similar to expectancy (e.g. “the trip made me hopeful”).

##### Post-efficacy Effect Isolation

We further discuss assumptions for post-efficacy effect isolation here. The pre-/post-efficacy expectancy definitions are critical for defining distinct sets of confounders. Confounders of the *E* → *Y* pathway may be caused by sets of confounders that are distinct before and after *t*^∗^ (i.e., pre/post-efficacy). By precisely defining these time intervals, we can reason about what these confounders might be, what statistical quantity to target, and how to implement modern causal inference techniques to estimate it. For example, say baseline (pre-randomization) expectancy is a reasonable proxy for the unmeasured confounders that drive *E* → *Y* before *t*^∗^. Would we expect baseline expectancy to also be the sole common cause of *E* → *Y* after *t*^∗^? Perhaps a larger set of confounders, including prior experience with psychedelics, depression severity, treatment history, and more might be necessary to estimate treatment effects not mediated through the unwanted expectancy pathways. Moreover, this emphasizes that the timing of the expectancy measurement is critical for both interpretation of mediation effects and the set of confounders that would need to be measured. Earlier measurements (e.g., not long after treatment administration) may represent a reasonable default in cases where immediate functional unmasking occurs, as fewer post-treatment confounders would need to be included. Measuring confounders of the post-efficacy *E* → *Y* pathway may be harder, but alternative experimental designs may provide a means to measure them. For example, experimenters might include a “lead-in” whereby all participants are treated with either an active comparator (e.g., THC for an psilocybin trial), or the actual treatment itself. The changes in expectancy to this initial lead-in treatment, in addition to other baseline covariates, might provide a proxy for the unmeasured confounders that drive post-efficacy *E* → *Y* paths.

#### E.2 Statistical Estimation

Recall that the causal effects of interest are composed of contrasts of average potential outcomes: *χ*(*a, e*):= 𝔼[*Y* (*a, e*)]. Under the assumptions outlined above, this effect can be estimated from the observed data distribution of the study, namely *χ*(*a, e*) = 𝔼(*h*^*a,e*^(*X*)), where

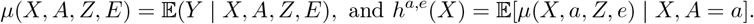

The proposed estimation strategy plugs-in predictions from four models into an equation to obtain an average treatment effect point estimate and 95% confidence intervals (CIs). By fitting these four models with flexible machine learning methods (e.g., random forest, neural networks), one avoids the need to specify a parametric relationship between the confounders and the outcome variable(s). This is desirable as parametric approaches, like including confounders as covariates in GLM, is only valid under strong assumptions. These four fitted models fall under “treatment models” and “outcome models” based on all causes (e.g., *X, Z*, and *A*).

The first “treatment model”, ℙ(*A* = *a* | *X*) is given by the known treatment randomization probabilities in a trial. That is, we can plug in the experimental treatment arm randomization probabilities ℙ(*A* = *a* | *X*) ≡ ℙ(*A* = *a*) when treatment is marginally randomized. Next, we model mean expectancy levels given its causes: baseline covariates, *X*, treatment arm, *A*, and post-treatment confounders of the *E* − *Y* effect, *Z*. We denote this as *π*_1_(*e* | *X, A, Z*):= ℙ[*E* = *e* | *X, A, Z*]. If *E* is binary, this regression might be, for example, a logistic regression model of expectancy, *E*, using *A, E*, and *Z* as features/covariates, though in practice we use flexible supervised learning methods.

We next fit two “outcome models” of the conditional mean *Y*, given its causes. The first is the conditional expectation *µ*(*X, A, Z, E*):= 𝔼[*Y* | *X, A, Z, E*]. For example, if *Y* is continuous, one could use a linear regression using *A, X, E*, and *Z* as covariates, though again, in practice we use flexible methods. Then this model’s predictions on study data, 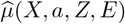, at fixed levels of treatment and expectancy, are used as the outcome variable for the secondary outcome model 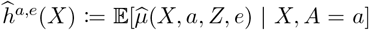. Finally, we calculate the causal effect estimates, and obtain confidence intervals and p-values, by plugging in predictions from these models into closed-form equations to get estimates of the CDE.

Our implementation involves additional steps to improve the statistical properties of this second outcome model (Díaz et al., 2023). In practice, we suggest an ensemble of flexible machine learning methods for all of the above “nuisance” models. We use cross-fitting, a crossvalidation like procedure to avoid an overfitting-related bias (Chernozhukov et al., 2018). This splits the dataset into non-overlapping model-fitting and model-prediction folds. We use each fold to derive an estimate, and then combine the estimates across folds, and calculate a pooled variance estimate to obtain confidence intervals and p-values. In the appendix, we provide code, and a step-by-step guide for implementing such analyses through an adaptation of the methods implemented in the lmtp package (Williams and Díaz, 2023; Díaz et al., 2021). These methods are often known as “doubly-robust”, “double ML”, and shares close connections with “targeted minimum loss estimation” methods which we recommend as an appropriate alternative (see comparison in Liu et al. (2016)).

The strategy requires sufficient sample sizes, as theoretical guarantees are asymptotic, but it is difficult to offer specific guidance without strong assumptions about effect sizes. The approach is, however, applicable to cases where expectancies are measured at multiple timepoints, and/or participant dropout leads to missing data (Liu et al., 2016).

#### E.3 Comparison with other statistical strategies for functional unmasking in the psychedelic literature

Additional mediation approaches have been proposed to account for functional unmasking in psychedelic RCTs. Muthukumaraswamy et al. (2021) proposed mediation analyses using parametric models that make strong assumptions about the functional form of the belief-outcome relationship, that we think are unrealistic in practice. In our view, they rightly conceptualize failure of blinding as an individual-level effect of treatment on belief, and recognize conditioning on post-treatment belief as a source of bias. However, they propose to base policy on a statistical quantity (and associated hypothesis tests) using specific so-called *principal stratum* effects (see expression 14 in Muthukumaraswamy et al. (2021)), which we caution against in general (see Stensrud et al. (2023) for a detailed discussion). Briefly, these effects are interpreted as the causal effect of treatment among participants who would have the same level of belief in both treatment arms (i.e. the people who *would be* successfully blinded). This is a treatment effect in a subset of the population that can never be observed because it is defined by a post-treatment status within the same participants under *different* treatments (e.g. placebo vs. psilocybin). In fact, this subset of the population may not exist, and even when it does, it may be highly idiosyncratic. For example, this statistical quantity may describe the causal effect of psilocybin among people for whom psilocybin has little therapeutic benefit, thereby explaining why they meet the principal stratum definition that psilocybin administration would result in the same level of belief/expectancy in treatment and placebo arms. Finally, this approach rests on assumptions that are not falsifiable in any experiment. Taken together, we recommend against using this strategy for policy making.

Next, use of biomarkers in mediation analyses (Muthukumaraswamy, 2023) is intuitively appealing, and our framework provides theoretical context that can be used to reason about how to apply such a strategy. We believe a more statistically modern version of this (akin to the the strategy described above for mediation by *E*) that does not make such strong parametric assumptions would be a fruitful way to answer questions related to a specific biological mechanism of action of psychedelics. It may, however, be more difficult to use as a method to handle functional unmasking. If the goal is to understand the role of a biological process, we recommend considering whether the process takes place pre-/-post-efficacy to clarify 1) what confounders must be measured, and 2) what effects the analysis isolates. For example, if the biomarker is striatal BOLD activity measured during the trip, and one believes the therapeutic benefit of psilocybin is neurogenesis that takes place a week after the trip, then the biomarker may serve as a pre-efficacy mediator. One would have to then measure all common causes of that BOLD activity and the outcome and adjust for them in properly during estimation. On the other hand, if the mediator (e.g. BOLD activity) is used as an “objective” measure of the “biological mechanism” responsible for the therapeutic effects of the compound, then conducting a mediation analysis of the biomarker may not resolve the functional unmasking problem when it causes future expectancies. For example, estimating an indirect effect of the treatment *A*, mediated through the biomarker, *M*, (e.g., through the treatment-fixed CDE of the mediator 𝔼[*Y* (*a, m*) − *Y* (*a, m*^′^)]), does not result in post-efficacy isolated effects if say, post-efficacy expectancy, *E*_*t*_∗, is mediated through a *post-efficacy* mediator. In sum, biomarker mediation analyses must apply the same reasoning as expectancy measurements described above, about what potential confounders are, and what the interpretation of the resulting causal effect estimate is (e.g., is the biomarker measured pre/post-efficacy).

Some have suggested comparing open label and RCT studies to estimate the magnitude of expectancy effects (Szigeti et al., 2024a). This is only valid if one properly adjusts for all common causes of study participation and the outcome, as the populations may differ across studies. If one believes they have measured all potential confounders, then one can apply methods from the rich causal transportability literature (Cole and Stuart, 2010; Degtiar and Rose, 2023; Vuong et al., 2025) and negative control causal inference literature (Lipsitch et al., 2010; Tchetgen Tchetgen et al., 2024) to formally combine data from different sources and adjust for confounding. Otherwise, informal strategies (Szigeti et al., 2024a) could in principle result in bias so strong that it makes beneficial treatments appear harmful, or vice-versa.

We commend Muthukumaraswamy et al. (2025) for highlighting potential drawbacks of RCTs in this setting. Although we do not agree with all arguments presented, we agree that “real world evidence” and observational studies in psychedelics can teach us a lot and require fewer resources. Flexible tools from modern causal inference will be critical in drawing inferences without strong assumptions in these observational settings. Our example code can be easily adapted to estimate, for example, average treatment effects in observational studies (with or without expectancy measurements). This conceptualization might allow researchers to explore how to flexibly incorporate measured qualities of the “trip” (e.g., set, setting) and baseline participant characteristics to account for expectancy.

#### E.4 Problems with Alternative Causal Strategies

##### E.4.1 Correct Guess Rate Curve

Szigeti et al. (2023) propose the so-called *Correct Guess Rate Curve*, and claim that it is a statistical tool that can “estimate the outcome of a perfectly blinded trial based on data from an imperfectly blinded trial.” The implied estimand that is targeted by their approach is not explicitly formalized; in this section we derive this estimand and show that it does not, in general, satisfy the properties claimed in Szigeti et al. (2023).

The proposed approach uses kernel density estimation to fit the *distribution* of the outcome in the four treatment-belief strata, then repeatedly draws from these four distributions in pre-specified proportions to construct pseudo-populations on which they conduct a “corrected” analysis. For the purposes of estimating average outcomes in the hypothetical “perfectly blinded trial”, the approach ultimately boils down to comparisons of weighted sums of the means in these four strata,

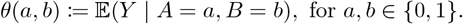

The authors draw half of their samples from the “correct guess” strata, [*A* = 0, *B* = 0] and [*A* = 1, *B* = 1], such that “the relative sample sizes of the correct guess strata are preserved.” Mathematically, this means that the strata [*A* = 0, *B* = 0] and [*A* = 1, *B* = 1] represent 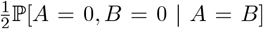 and 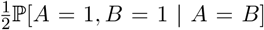 of the total constructed pseudo-population, respectively. Similarly, the other half of samples are drawn from the “incorrect guess” strata, [*A* = 0, *B* = 1] and [*A* = 1, *B* = 0], again retaining the same relative proportion as in the original data. This likewise means that the strata [*A* = 0, *B* = 1] and [*A* = 1, *B* = 0] represent 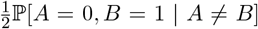 and 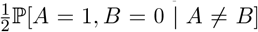 of the total constructed pseudo-population, respectively. As a result of this process, the authors propose to “estimate the direct treatment effect” by contrasting treated versus untreated in the constructed pseudo-populations. Notice that, using 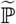 to denote the data distribution in the pseudo-population,

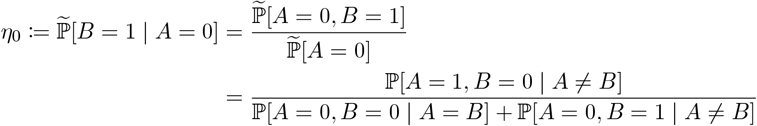

and

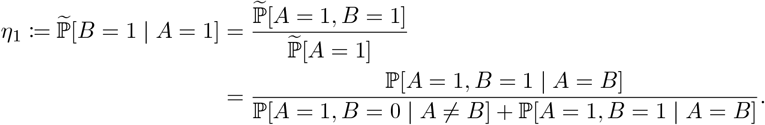

Moreover, the treatment effect in the pseudo-population is given by

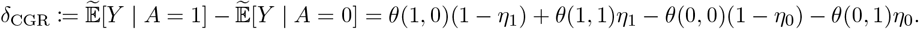

In general, this estimand will not have a clear causal interpretation, nor will it represent a “direct effect” of treatment.

To illustrate, suppose treatment is randomly assigned as *A* ∼ Bernoulli(1*/*2), and *U* ∼ Unif(0, 1) represents an unmeasured common cause of belief and the outcome. Suppose further that beliefs are generated according to

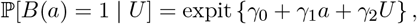

and outcomes satisfy the generative model

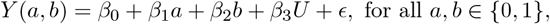

where *ϵ* is a mean-zero auxiliary noise variable. Then the individual level controlled direct effect is *Y* (1, *b*) − *Y* (0, *b*) = *β*_1_, for each individual in the population. On the other hand,

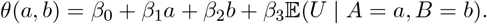

Obtaining *δ*_CGR_ by combining these with the implied weights for *η*_*a*_ will not, in general, recover *β*_1_. For concreteness, taking the following parameter specification:

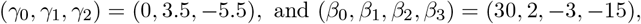

we can show that *δ*_CGR_ ≈ − 0.38, whereas the CDE is *β*_1_ = 2. In this case, one would erroneously conclude a harmful effect of treatment (assuming greater *Y* values are favorable) when in fact the treatment is beneficial. While the coefficients on *U* in this specification are set to large values for illustration purposes, bias occurs for smaller coefficient values as well.

##### E.4.2 Belief-Conditional Effects

In the main text, we described how an analysis presented at the MDMA Lykos trial FDA panel committee discussion, as well as other studies on psychedelics (Bershad et al., 2019; van Elk et al., 2022; Lii et al., 2023a; Cavanna et al., 2022), have attempted to assess treatment effects conditional on post-treatment belief (e.g., see Figure 1). We appreciate that these analyses are sometimes conducted in an exploratory fashion, but we wish to illustrate here why this approach can lead to biased results and highly misleading conclusions.

Mathematically, using the notation from the previous section, this approach centers on the contrast

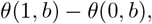

where *b* is chosen to be 0 (i.e., those that believe they received the control condition) or 1 (i.e., those that believe they received the active treatment). Similar to the correct guess rate curve estimand, this contrast does not recover any kind of direct effect in general. Taking the same generative scenario from the previous section, we find that

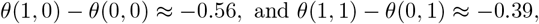

when the true CDE is 2 for each individual.

In the manuscript, Figure 1, we reformulated the problem to instead plot changes in the outcome from baseline (pre-randomization) so that that the plot was more comparable to the MDMA Lykos trial FDA panel committee discussion slide (e.g., was on a similar scale). Without loss of generality, we assume each participants’ baseline score was normalized to be 0. Specifically, we took the following parameter specification:

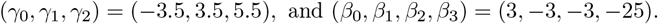

#### E.5 Causal Effects when Experimental Manipulation of Expectancy is Incomplete

There is disagreement on the feasibility of completely manipulating expectancy (Szigeti and Heifets, 2024; Bingel et al., 2011). If expectancy is not fully controlled by experimental manipulation (in either arm), the treatment effect may still be partially mediated through *E*. In particular, if the sensitivity of expectancy levels to expectancy manipulations differs substantially between-arms, the CDE may have similar “contamination” as the ATE. Nevertheless, even when expectancy cannot be completely controlled experimentally, the joint impact of treatment and expectancy manipulations can help formally probe the impact of unmasking by testing the sensitivity of the outcome to changes in expectancy levels.

One could, in principle, use instrumental variable methods to account for differing effectiveness of messages across treatment arms. By conceptualizing expectancy manipulation interventions as “instruments”, and the subsequently measured expectancy level as a “treatment”, one can apply these methods to estimate treatment effects in terms of expectancy, itself, rather than the message intervention. Under certain conditions, this strategy can yield an appealing causal effect interpretation, and account for differing responsiveness or “compliance” to messages across individuals. The application of this strategy typically requires one to make one of two sets of unverifiable assumptions. For example, “homogeneity assumptions” require that any unmeasured common causes of expectancy and *Y* do not modify the strength of the *E*_*t*_ → *Y* path. This is unrealistic: for instance, homogeneity would imply that the therapist-participant interactions would not alter how expectancies drive the outcome. Alternatively, one can rely on “monotonicity” assumptions (Hernan and Robins, 2023). These are often deemed justifiable, but only allow one to identify (i.e. result in a valid causal effect that can be estimated from study data) effects of expectancy *within* (but not across) treatment arms, *A*, in a latent (unobserved) subgroup. Critically, this would not allow one to identify the causal quantity of interest: a controlled direct effect of the psychedelic at a fixed level of expectancy. We also wish to acknowledge that more broadly, there have been past proposals for the use of instrumental variable methods in unblinded settings (Chaibub Neto, 2016).

#### E.6 Estimands for Sequentially Randomized Experiments

We now describe statistical quantities that can be estimated 1) non-parametrically as simple means within subgroups of treatment arms, or 2) semi-parametrically with a MSM. These statistical summaries, or “estimands” allow us to test questions relating to timing, durability, expectancy, and treatment dose. For ease of exposition, we focus on the simple case of a two timepoint study, but note that these are easily generalizable to more than two timepoints. We cast all estimands in terms of the outcome counterfactual *Y* ((*m*_1_, *m*_2_), (*a*_1_, *a*_2_)): the outcome variable that would be observed if a participant got the expectancy manipulation message, *m*_1_, and treatment *a*_1_ at timepoint 1, and the message, *m*_2_, and treatment *a*_2_ at timepoint 2. We also express some in terms of expectancy counterfactuals *E*_*j*_ ((*m*_1_, *m*_2_), (*a*_1_, *a*_2_)): the expectancy that would be observed under the treatment sequences (*m*_1_, *m*_2_), (*a*_1_, *a*_2_). Population means of these counterfactuals represent quantities of interest in their own right. Moreover, contrasts of these quantities yield various effects with important interpretations, and corresponding null hypothesis tests (for the contrast equaling some null value):

- Time-dependency Effects of Messages: 𝔼[*Y* ((1, 0), (*a*_1_, *a*_2_))] − 𝔼[*Y* ((0, 1), (*a*_1_, *a*_2_))]
- Controlled Dose Effect of Message: 𝔼[*Y* ((0, 0), (*a*_1_, *a*_2_))] − 𝔼[*Y* ((1, 1), (*a*_1_, *a*_2_))]
- “Message-Treatment Alignment”: 𝔼[*Y* ((0, 1), (1, 0))] − 𝔼[*Y* ((1, 0), (1, 0))]
- Effects of Treatment on Expectancy: 𝔼[*E*_*j*_((*m*_1_, *m*_2_), (0, 0))] − 𝔼[*E*_*j*_((*m*_1_, *m*_2_), (1, 1))]
- Controlled Direct Treatment Effects: 𝔼[*Y* ((*m*_1_, *m*_2_), (0, 0))] − 𝔼[*Y* ((*m*_1_, *m*_2_), (1, 1))]

The “Time-dependency Effects of Messages” tests the degree to which the recency of an expectancy manipulation affects the outcome. This might test the durability of expectancy effects, which helps answer: “if expectancy effects are a substantial portion of a treatment’s therapeutic benefit, how long does that expectancy-induced benefit last?” Similarly, one can test an expectancy dose-response effect with the “Controlled Dose Effect of Message” quantity: “do larger ‘doses’ of expectancy lead to greater therapeutic benefit?” To optimize treatment protocols, one can test the affect of aligning the treatment and the information (“messages”) provided to participants. This tests whether for a given “dose” of message, the degree to which the message is consistent with the actual treatment affects the outcome. This provides a timevarying way to test past finding that the success of expectancy manipulation interventions differs across treatment arms (Szigeti and Heifets, 2024). One can test the “Effects on Treatment on Expectancy” to identify the effect of a given treatment sequence since expectancy, itself, is a valid outcome. For example, if expectancy does not differ substantially between those receiving placebo or treatment, then one might conclude that expectancy is unlikely to be a major mediator of treatment effects. These estimands provide a formal testing framework to implement calls to measure expectancy throughout the study (Colloca et al., 2023; Szigeti and Heifets, 2024). Finally, one can use the “Controlled Direct Treatment Effects” described in the mediation section above to test how for a fixed expectancy manipulation dose, the treatment causes changes in the outcome.

As alluded to earlier, estimation of the individual counterfactual means is straightforward in designs where treatment and messages are sequentially randomized. However, power may be limited in testing equality certain combinations of parameters. As a potentially exploratory tool, one can adopt an MSM which places structure on the set of causal quantities:

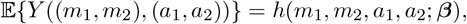

for some function *h* parametrized by ***β***. For example, if dose and messaging are thought to be additive in nature, one might adopt

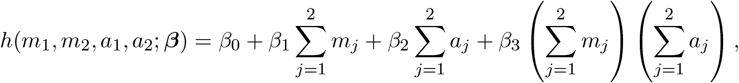

where ***β*** = (*β*_0_, *β*_1_, *β*_2_, *β*_3_). In this sequentially randomized setting, estimates of these MSM parameters can be obtained by regressions, with robust variance estimators applied to calculate confidence intervals and p-values.

#### E.7 Ideal Active Controls (“Deep blinding”)

With the aim of attaining isolated effects through well thought-out study design, our formal counterfactual framework suggests that the ideal active comparator should 1) cause comparable expectancy levels as the treatment, and 2) allow for expectancy manipulations to have similar effectiveness on expectancy levels as the active treatment. It is instructive to consider how different active controls operate in terms of potential belief blinding and/or expectancy blinding. One natural goal is to seek a control that has identical characteristics as the treatment but without the therapeutic properties. A successful choice would result in identical post-treatment beliefs and expectancies whereby a contrast of average outcomes between-arms would inherently represent a direct of effect of treatment not mediated through expectancy. This approach may, however, fail or be ill-defined in psychedelic studies that use psychoactive active comparators (e.g. methylphenidate for MDMA, or THC for psilocybin). For example, would the active comparator ideally cause “the same trip but without the insight”? One might argue that any active comparator is satisfactory as long as it results in belief blinding: a participant is unable to guess what compound they received. We emphasize, however, that belief-blinding is not always sufficient to control expectancy effects. Consider the example described in main text Section, in which salvia is used as an active comparator for smoke-able DMT treatment. If the sample of participants has no psychedelic experience or knowledge, then the strong psychoactive effects of salvia may result in an inability of participants to guess their treatment assignment. Nevertheless, the dysphoric properties of salvia may result in salvia arm participants reporting far worse expectancies than if they had instead smoked an inactive placebo. Thus even if DMT had no therapeutic benefits beyond expectancy effects, the treatment effect estimate may be large in this study only because salvia led to very negative expectancy levels.

Thus, we propose that an active comparator should ideally result in the same expectancy levels, *E*_*t*_, to the treatment across *pre-efficacy* timepoints: *E*_*t*_(*a* = 1) = *E*_*t*_(*a* = 0) for all timepoints, *t* = 1, 2, …, *t*^∗^ − 1. Moreover, we believe that a good active comparator would result in expectancy levels that can be experimentally manipulated (through a message *M*) to the same degree across treatment arms: *E*_*t*_(*m* = 1, *a* = 0) − *E*_*t*_(*m* = 0, *a* = 0) = − *E*_*t*_(*m* = 1, *a* = 1) − *E*_*t*_(*m* = 0, *a* = 1) for all *t < t*^∗^. This is critical because if expectancy cannot be manipulated in a comparable manner across treatment arms, even experimental expectancy manipulation may not control functional unmasking. This “interventionist” conceptualization provides hypothesis tests and possible interventions to evaluate the effectiveness of an active comparator. In sum, compared to the treatment, the ideal active comparator should result in expectancy levels that are comparable across treatment arms, and can be experimentally manipulated to the same degree.

### F Controlled Direct Effects Code Guide

#### Controlled Direct Effects for Unmasking

**Gabriel Loewinger, Mats Stensrud, Alex Levis**

**2025-09-25**

#### lmtp R package

We use the lmtp R package. In addition to the two papers in the references section below, the following resources may also be helpful: - 1) https://muse.jhu.edu/pub/56/article/883479/pdf (https://muse.jhu.edu/pub/56/article/883479/pdf) - 2) https://beyondtheate.com/ (https://beyondtheate.com/)

#### Data Formating

Let’s load the data, look at the format, and work through a simple implementation. We simulated this data for demonstration purposes. The code we used to simulate the data is available on our Github repo, although the details are less important than understanding the steps in the analysis code below so one can apply a similar analysis on a real dataset.

~~~
dat <-read.csv(“CDE_data.csv”)
head(dat)
~~~

|  | A | E | B | SS | mf | age | psych_h | Y | id |
| --- | --- | --- | --- | --- | --- | --- | --- | --- | --- |
|  | <int> | <int> | <int> | <dbl> | <int> | <dbl> | <dbl> | <dbl> | <int> |
| 1 | 0 | 1 | 1 | 0.04211587 | 0 | 0.94 | 0.32877732 | 4.677747538 | 1 |
| 2 | 0 | 0 | 0 | -0.91092165 | 1 | 0.62 | 0.45313145 | -0.005458508 | 2 |
| 3 | 1 | 0 | 1 | 0.15802877 | 0 | 0.70 | 0.50044097 | 6.575391876 | 3 |
| 4 | 1 | 0 | 0 | -0.65458464 | 0 | 0.40 | 0.18086636 | 4.441495292 | 4 |
| 5 | 0 | 0 | 1 | 1.76728727 | 0 | 0.64 | 0.52963060 | 2.853519677 | 5 |
| 6 | 1 | 1 | 1 | 0.71670748 | 0 | 0.80 | 0.07527575 | 6.596542880 | 6 |
| 6 rows |  |  |  |  |  |  |  |  |  |

~~~
dat-as.data.frame(dat) *# set to data.frame*
~~~

Each row is a participant (*n* = 50). The columns contain the following variables:

- A is a binary indicator of treatment/control
- E is the post-treatment Expectancy measurement
- B is the post-treatment Belief measurement
- SS is a set/setting variable measured during treatment administration
- mf is male/female biological sex
- age is participant age
- psych_h is psychedelic history/experience prior to the study
- Y is the outcome (e.g., depression levels)
- id is the subject ID

#### Controlled Direct Effect

We use the counterfactual notation *Y* (*a, e*) to be the outcome value that would be observed for a given participant in a world where we set their treatment value *A* = *a* and expectancy level *E* = *e*.

The goal of the analysis is to formalize and answer the question ``what is the difference in the outcome variable *Y* (e.g., depression levels) at follow-up among those treated with psilocybin *a* = 1 or control *a* = 0, in a world where we set pre-efficacy expectancy levels, *E* to the same value *E* = *e* across treatment arms?’’ This is the so-called controlled direct effect:

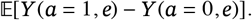

We show code for the simple setting where *E* is binary and indicates whether one does (*E* = 1) or does not (*E* = 0) expect that the treatment (that they think they received) will improve their outcome variable (e.g., Depression) at follow-up.

#### Possible treatment sequences

There are four possible treatment sequences (“regimes”/”policies”) of (*a, e*): (1,1), (1,0), (0,1), (0,0)). We can use the two pairs of these sequences to form contrasts of the causal effect of treatment when we “set” post-treatment expectancy at either *e* = 0 or *e* = 1. We have to code this as four separate functions which we do below:

~~~
*# A == 1, E == 0*
d_A1_E0 <-**function**(data, trt) {
     rep( ifelse(trt == “A”, 1, 0),
        nrow(data)
       )
}
*# A == 0, E == 1*
d_A0_E1 <-**function**(data, trt) {
     rep( ifelse(trt == “E”, 1, 0),
        nrow(data)
    )
     }
*# A == 1, E == 1*
d_A1_E1 <-**function**(data, trt) {
     **return**( rep(1, nrow(data)))
};
*# A == 0, E == 0*
d_A0_E0 <-**function**(data, trt) {
     **return**( rep(0, nrow(data)))
};
~~~

#### Confounders

Now we need to include the vectors of confounders *Z* of the *E* → *Y* path. In the DAG in the paper, we used the vector *X* to denote baseline variables (pre-randomization). Here we assume that together the vector of variables ( *A, B, X, Z*) are the complete set of common causes of *E* → *Y* path. We will distinguish between *X* and *Z* in the code below for coding convenience, but in reality, the estimation function below uses all variables (*A, B, X, Z*) appropriately.

First let’s specify the variable names in the dataset associated with the baseline and post-treatment variables (remember these are *all* confounders of *E* → *Y*), we only distinguish between them here for coding purposes.

~~~
X <-c(“psych_h”, “age”, “mf”) *# baseline variables*
Z <-c(“SS”, “B”) *# post-treatment confounders*
~~~

#### Estimation of CDE

To target a controlled direct effect (CDE), we can pretend “A” and “E” are longitudinal treatments, which we specify in the trt argument. We specify the name of the outcome variable, the type (here it is continuous but this package also handles survival and binary outcome types). Since treatment *A* is randomized at baseline, we can specify baseline = NULL. In the confounders list time_vary, we specify that it is a list that has two elements, the first list contains a vector of variable names that are confounders for *A* → *Y* and the second for *E* → *Y*. Because we set the variable k = Inf, these variables are pooled across timepoints, so the lmtp_sdr function treats all the variables in *X* and *Z* (concatenated together) as the confounders of the *E* → *Y* pathway. Since *A* is randomized, we can either set the known treatment randomization probabilities, or estimate them using the baseline variables in *X*. Both are valid, but here we show the case where these are estimated.

We set the shift variable to be the relevant treatment sequence that we set above. Remember we are interested in the two sequences where *e* = 0. mtp should be set to FALSE (see package documentation for details), and the participant id variable is set to the relevant column name in the dataset. The folds argument is the number of folds used in cross-fitting, a cross-validation like procedure used to estimate the different nuisance functions. If the dataset is very small (e.g., *n* <= 75), we think it makes sense to set folds = 1. Otherwise, we recommend setting folds = 2. The learners_trt and learners_outcome are the machine learning prediction models. We used a GLM and a XGBoost here, but there are many options that can be used. See SuperLearner R package for more information on the names. Whatever learners that we use (e.g., XGBoost, random forest), make sure you install that R package first. In practice, setting 2-4 learners is probably sufficient for our purposes. We recommend including a GLM as one of them, especially since sample sizes are likely to be small-moderate in these applications, and the confounders are likely to be low dimensional (e.g., regularization with Ridge/LASSO is unlikely to be as important as in many other settings where the confounders may be high dimensional). Including some algorithms that are flexible (e.g., GAMs, Random Forest, XGBoost) may help capture non-linearities and interactions among confounders. As an example, we show how to install the packages needed for this Vignette here:

~~~
install.packages(“lmtp”)
install.packages(“xgboost”)
~~~

#### Low Expectancy (*e* = 0)

Now let’s fit two functions for the two combinations needed for the contrast:

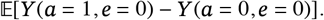

~~~
*# load packages*
**library**(lmtp)
**library**(xgboost)
*# d_A1_E0*
fit_sdr_A1E0 <-lmtp::lmtp_sdr(
    data = dat,
    trt = c(“A”, “E”), *# second timepoint’s “treatment”/exposure is expectancy, E*
    outcome = “Y”,
    outcome_type = “continuous”,
    baseline = NULL, *# no time-varying confounders: Baseline confounders are always includ*
ed in Ht according to documentation
    time_vary = list(X, Z), *# confounders of E -> Y path*
    k = Inf, *# whether to use all previous history variables pooled/concatenated*
    shift = d_A1_E0,
    id = “id”,
    mtp = FALSE,
    folds = 2,
    learners_trt = c(“SL.glm”, “SL.xgboost”, “SL.mean”),
    learners_outcome = c(“SL.xgboost”, “SL.glm”)
)
*# d_A0_E0*
fit_sdr_A0E0 <-lmtp::lmtp_sdr(
    data = dat,
    trt = c(“A”, “E”), *# second timepoint’s “treatment”/exposure is expectancy, E*
    outcome = “Y”,
    outcome_type = “continuous”,
    baseline = NULL, *# no time-varying confounders: Baseline confounders are always includ*
ed in Ht according to documentation
    time_vary = list(X, Z), *# confounders of E -> Y path*
    k = Inf, *# whether to use all previous history variables pooled/concatenated*
    shift = d_A0_E0,
    id = “id”,
    mtp = FALSE,
    folds = 2,
    learners_trt = c(“SL.glm”, “SL.xgboost”, “SL.mean”),
    learners_outcome = c(“SL.xgboost”, “SL.glm”)
)
~~~

Now that we have fit the above to models, we can extract a contrast with a point-estimate, confidence intervals and a p-value. We use the function below to compare the two fitted estimates, specifying the reference (i.e., the control and low expectancy (*a* = 0, *e* = 0) combination). We set the contrast to be on the additive scale, but there may be cases where one might wish to use a relative risk or other contrast type.

~~~
lmtp::lmtp_contrast(fit_sdr_A1E0, ref = fit_sdr_A0E0, type = “additive”)
## LMTP Contrast: additive
## Null hypothesis: theta == 0
##
## shift ref estimate std.error conf.low conf.high p.value
## 1 5.03 2.15 2.88 0.29 2.31 3.44 <0.001
~~~

In this dataset, we simulated the true *CDE* = 3. We can see the confidence intervals contain the true value and the point estimate is reasonably close to the truth.

#### High Expectancy (*e* = 1)

Now let’s fit two functions for the two combinations needed for the contrast:

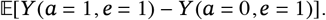

Now let’s repeat these same steps with the two other relevant policies for the *e* = 1 (high expectancy) setting.

~~~
*# E == 1
# d_A1_E1*
fit_sdr_A1E1 <-lmtp::lmtp_sdr(
     data = dat,
     trt = c(“A”, “E”), *# second timepoint’s “treat     ment”/exposure is expectancy, E*
     outcome = “Y”,
     outcome_type = “continuous”,
     baseline = NULL, *# no time-varying confounders: Baseline confounders are always included in Ht according to documentation*
     time_vary = list(X, Z), *# confounders of E -> Y path*
     k = Inf, *# whether to use all previous history variables pooled/concatenated*
     shift = d_A1_E1,
     id = “id”,
     mtp = FALSE,
     folds = 2,
     learners_trt = c(“SL.glm”, “SL.xgboost”, “SL.mean”),
     learners_outcome = c(“SL.xgboost”, “SL.glm”)
)
*# d_A0_E1*
fit_sdr_A0E1 <-lmtp::lmtp_sdr(
     data = dat,
     trt = c(“A”, “E”), *# second timepoint’s “treatment”/exposure is expectancy, E*
     outcome = “Y”,
     outcome_type = “continuous”,
     baseline = NULL, *# no time-varying confounders: Baseline confounders are always included in Ht according to documentation*
     time_vary = list(X, Z), *# confounders of E -> Y path*
     k = Inf, *# whether to use all previous history variables pooled/concatenated*
     shift = d_A0_E1,
     id = “id”,
     mtp = FALSE,
     folds = 2,
     learners_trt = c(“SL.glm”, “SL.xgboost”, “SL.mean”),
     learners_outcome = c(“SL.xgboost”, “SL.glm”)
)
lmtp::lmtp_contrast(fit_sdr_A1E1, ref = fit_sdr_A0E1, type = “additive”)
##
## shift ref estimate std.error conf.low conf.high p.value
## 1 5.53 2.62 2.91 0.273 2.38 3.45 <0.001
Again we can see the high expectancy contrast yields a value close to the true *CDE* = 3.
~~~

#### How to Cite

For use of this code or vignette, please cite the following papers:

Gabriel Loewinger∗, Mats Stensrud, David Yaden, Alex Levis. A Causal Inference Framework for Effect Isolation in Randomized Studies with Unmasking (https://arxiv.org/abs/2405.18597v1). biorXiv (2025).

Williams, N. and Díaz, I. (2023), “lmtp: an R package for estimating the causal effects of modified treatment policies,” Observational Studies, 9, 103–122

Díaz, I., Williams, N., Hoffman, K. L., and Schenck, E. J. (2023), “Nonparametric causal effects based on longitudinal modified treatment policies,” Journal of the American Statistical Association, 118, 846–857.

## References

Aday, J. S., Heifets, B. D., Pratscher, S. D., Bradley, E., Rosen, R., and Woolley, J. D. (2022), “Great expectations: recommendations for improving the methodological rigor of psychedelic clinical trials,” Psychopharmacology, 239, 1989–2010.

Barrett, F. S., Carbonaro, T. M., Hurwitz, E., Johnson, M. W., and Griffiths, R. R. (2018), “Double-blind comparison of the two hallucinogens psilocybin and dextromethorphan: effects on cognition,” Psychopharmacology, 235, 2915–2927.

Bershad, A. K., Mayo, L. M., Van Hedger, K., McGlone, F., Walker, S. C., and de Wit, H. (2019), “Effects of MDMA on attention to positive social cues and pleasantness of affective touch,” Neuropsychopharmacology, 44, 1698–1705.

Bingel, U., Wanigasekera, V., Wiech, K., Ni Mhuircheartaigh, R., Lee, M. C., Ploner, M., and Tracey, I. (2011), “The effect of treatment expectation on drug efficacy: imaging the analgesic benefit of the opioid remifentanil,” Science translational medicine, 3, 70ra14–70ra14.

Butler, M., Jelen, L., and Rucker, J. (2022), “Expectancy in placebo-controlled trials of psychedelics: if so, so what?” Psychopharmacology, 239, 3047–3055.

Cavanna, F., Muller, S., de la Fuente, L.A., Zamberlan, F., Palmucci, M., Janeckova, L., Kuchar, M., Pallavicini, C., and Tagliazucchi, E. (2022), “Microdosing with psilocybin mushrooms: a double-blind placebo-controlled study,” Translational Psychiatry, 12, 307.

Chaibub Neto, E. (2016), “Using instrumental variables to disentangle treatment and placebo effects in blinded and unblinded randomized clinical trials influenced by unmeasured confounders,” Scientific Reports, 6, 37154.

Chernozhukov, V., Chetverikov, D., Demirer, M., Duflo, E., Hansen, C., Newey, W., and Robins, J. (2018), “Double/debiased machine learning for treatment and structural parameters,” The Econometrics Journal, C1–C68.

Cheung, K., Propes, C., Jacobs, E., Earp, B. D., and Yaden, D. B. (2025), “Response to the US Congress Request for Information (RFI) on Psychedelic Research for Mental Health,” Submitted response, Hub at Oxford for Psychedelic Ethics, as submitted by the Hub at Oxford for Psychedelic Ethics.

Cole, S. R. and Stuart, E. A. (2010), “Generalizing evidence from randomized clinical trials to target populations: the ACTG 320 trial,” American Journal of Epidemiology, 172, 107–115.

Colloca, L. and Fava, M. (2024), “What should constitute a control condition in psychedelic drug trials?” Nature Mental Health, 2, 1152–1160.

Colloca, L., Nikayin, S., and Sanacora, G. (2023), “The intricate interaction between expectations and therapeutic outcomes of psychedelic agents,” JAMA psychiatry, 80, 867–868.

Degtiar, I. and Rose, S. (2023), “A review of generalizability and transportability,” Annual Review of Statistics and Its Application, 10, 501–524.

Devilly, G. J. and Borkovec, T. D. (2000), “Psychometric properties of the credibility/expectancy questionnaire,” Journal of behavior therapy and experimental psychiatry, 31, 73–86.

Díaz, I. (2024), “Non-agency interventions for causal mediation in the presence of intermediate confounding,” Journal of the Royal Statistical Society Series B: Statistical Methodology, 86, 435–460.

Dickerman, B. A., García-Albéniz, X., Logan, R. W., Denaxas, S., and Hernán, M. A. (2019), “Avoidable flaws in observational analyses: an application to statins and cancer,” Nature medicine, 25, 1601–1606.

Dickerman, B. A., Gerlovin, H., Madenci, A. L., Kurgansky, K. E., Ferolito, B. R., Muñiz, M. J. F., Gagnon, D. R., Gaziano, J. M., Cho, K., Casas, J. P., and Hernán, M. A. (2022), “Comparative Effectiveness of BNT162b2 and mRNA-1273 Vaccines in U.S. Veterans,” New England Journal of Medicine, 386, 105–115.

Díaz, I., Williams, N., Hoffman, K. L., and Schenck, E. J. (2021), “Non-parametric causal effects based on longitudinal modified treatment policies,” Journal of the American Statistical Association.

Gabriel, E. E., Ocampo, A., and Sjölander, A. (2025), “Elucidating some common biases in randomized controlled trials using directed acyclic graphs,” European Journal of Epidemiology, 1–11.

Goodwin, G. M., Aaronson, S. T., Alvarez, O., Arden, P. C., Baker, A., Bennett, J. C., Bird, C., Blom, R. E., Brennan, C., Brusch, D., et al. (2022), “Single-dose psilocybin for a treatment-resistant episode of major depression,” New England Journal of Medicine, 387, 1637–1648.

Hernan, M. and Robins, J. (2023), Causal Inference: What If, Chapman & Hall/CRC Monographs on Statistics & Applied Probab, CRC Press.

(2024), Causal Inference: What If, Chapman & Hall/CRC Monographs on Statistics & Applied Probab, CRC Press.

Hernán, M. A., Alonso, A., Logan, R., Grodstein, F., Michels, K. B., Willett, W. C., Manson, J. E., and Robins, J. M. (2008), “Observational studies analyzed like randomized experiments: an application to postmenopausal hormone therapy and coronary heart disease,” Epidemiology, 19, 766–779.

Hernán, M. A., Brumback, B., and Robins, J. M. (2001), “Marginal structural models to estimate the joint causal effect of nonrandomized treatments,” Journal of the American Statistical Association, 96, 440–448.

Howick, J. H. (2011), The philosophy of evidence-based medicine, John Wiley & Sons.

Hubbard, A., Jamshidian, F., and Jewell, N. (2012), “Adjusting for perception and unmasking effects in longitudinal clinical trials,” The international journal of biostatistics, 8, 1–20.

Jamshidian, F., Hubbard, A. E., and Jewell, N. P. (2014), “Accounting for perception, placebo and unmasking effects in estimating treatment effects in randomised clinical trials,” Statistical methods in medical research, 23, 293–307.

Kaertner, L. S., Steinborn, M. B., Kettner, H., Spriggs, M. J., Roseman, L., Buchborn, T., Balaet, M., Timmermann, C., Erritzoe, D., and Carhart-Harris, R. L. (2021), “Positive expectations predict improved mental-health outcomes linked to psychedelic microdosing,” Scientific reports, 11, 1941.

Kahan, B. C., Hindley, J., Edwards, M., Cro, S., and Morris, T. P. (2024), “The estimands framework: a primer on the ICH E9 (R1) addendum,” bmj, 384.

King Baudouin Foundation (2022a), “2022 Rousseeuw Prize for Statistics Awarded to James M. Robins, Miguel A. Hernán, Andrea Rotnitzky, Thomas Richardson, and Eric J. Tchetgen Tchetgen,” https://www.rousseeuwprize.org/2022, topic: Causal Inference with application in Medicine and Public Health. Ceremony held at KU Leuven, Belgium, October 12, 2022.

King Baudouin Foundation (2022b), “First Rousseeuw Prize Awarded to Causal Inference: Long Description for Statisticians,” Press release, King Baudouin Foundation, detailed technical description of the 2022 Rousseeuw Prize awarded to James M. Robins, Miguel A. Hernán, Thomas Richardson, Andrea Rotnitzky, and Eric J. Tchetgen Tchetgen for groundbreaking methodological contributions to Causal Inference with applications in Medicine and Public Health.

Levis, A. W., Loewinger, G., and Pereira, F. (2024), “Causal inference in the closed-loop: Marginal structural models for sequential excursion effects,” Advances in Neural Information Processing Systems, 37, 109123–109151.

Lii, T. R., Smith, A. E., Flohr, J. R., Okada, R. L., Nyongesa, C. A., Cianfichi, L. J., Hack, L. M., Schatzberg, A. F., and Heifets, B. D. (2023a), “Randomized trial of ketamine masked by surgical anesthesia in patients with depression,” Nature Mental Health, 1, 876–886.

Lii, T. R., Smith, A. E., Flohr, J. R., Okada, R. L., Nyongesa, C. A., Cianfichi, L. J., Hack, L. M., Schatzberg, A. F., and Heifets, B. D. (2023b), “Randomized trial of ketamine masked by surgical anesthesia in patients with depression,” Nature Mental Health, 1, 876–886.

Lipsitch, M., Tchetgen, E. T., and Cohen, T. (2010), “Negative controls: a tool for detecting confounding and bias in observational studies,” Epidemiology, 21, 383–388.

Liu, W., Zhang, Z., Schroeder, R. J., Ho, M., Zhang, B., Long, C., Zhang, H., and Irony, T. Z. (2016), “Joint estimation of treatment and placebo effects in clinical trials with longitudinal blinding assessments,” Journal of the American Statistical Association, 111, 538–548.

Lynch, K. G., Cary, M., Gallop, R., and Ten Have, T. R. (2008), “Causal mediation analyses for randomized trials,” Health Services and Outcomes Research Methodology, 8, 57–76.

Matvey, M., Kelley, D. P., Bradley, E. R., Chiong, W., O’Donovan, A., and Woolley, J. (2025), “Modifying Informed Consent to Help Address Functional Unmasking in Psychedelic Clinical Trials,” JAMA Psychiatry, 82, 311–318.

Miles, C. H. (2023), “On the causal interpretation of randomised interventional indirect effects,” Journal of the Royal Statistical Society Series B: Statistical Methodology, 85, 1154–1172.

Muthukumaraswamy, S., Baggott, M., Schenberg, E. E., Repantis, D., Wolff, M., Forsyth, A., and Noorani, T. (2025), “Psychedelic Assisted Therapy as a Complex Intervention: Implications for clinical trial design,”.

Muthukumaraswamy, S. D. (2023), “Overcoming blinding confounds in psychedelic randomized controlled trials using biomarker driven causal mediation analysis,” Expert Review of Clinical Pharmacology, 16, 1163–1173.

Muthukumaraswamy, S. D., Forsyth, A., and Lumley, T. (2021), “Blinding and expectancy confounds in psychedelic randomized controlled trials,” Expert review of clinical pharmacology, 14, 1133–1152.

Nayak, S. M. and Zahid, Z. (2025), “Out of control: Blinding, dose response, and psychosocial controls in psychedelic trials,” Journal of Psychopharmacology, 02698811251368367.

Obolski, U., Stensrud, M. J., and Nevo, D. (2024), “A Call for Blinding Assessments in Dengue Vaccine Trials,” The Lancet Infectious Diseases, 24, e10.

Ohlsson, H. and Kendler, K. S. (2020), “Applying causal inference methods in psychiatric epidemiology: A review,” JAMA psychiatry, 77, 637–644.

Organization, N. P. (2021), “The Prize in Economic Sciences 2021,” NobelPrize.org, the prize was awarded to Joshua Angrist and Guido Imbens for their methodological contributions to the analysis of causal relationships.

Pearl, J. (2001), “Direct and indirect effects,” in Proceedings of the 17th Conference on Uncertainty in Artificial Intelligence, 2001, San Francisco, CA: Morgan Kaufmann, pp. 411–420.

Pearl, J. (2011), “A.M. Turing Award,” [https://amturing.acm.org/awardwinners/pearl2658896.cfmACMA.M.Turi

Reagan-Udall Foundation for the FDA (2024), “Advancing Psychedelic Clinical Study Design: Meeting Summary,” Reagan-Udall Foundation for the FDA.

Richardson, T. S. and Robins, J. M. (2013), “Single world intervention graphs (SWIGs): A unification of the counterfactual and graphical approaches to causality,” Center for the Statistics and the Social Sciences, University of Washington Series. Working Paper, 128, 2013.

Robins, J. (1986), “A new approach to causal inference in mortality studies with a sustained exposure period—application to control of the healthy worker survivor effect,” Mathematical Modelling, 7, 1393–1512.

Robins, J. (1987), “A graphical approach to the identification and estimation of causal parameters in mortality studies with sustained exposure periods,” Journal of chronic diseases, 40, 139S–161S.

Robins, J. (1999), “Marginal Structural Models,” Proc. of ASA Sect. on Bayes. Stat. Sci., 1–10.

Robins, J. M. and Greenland, S. (1992), “Identifiability and exchangeability for direct and indirect effects,” Epidemiology, 3, 143–155.

Robins, J. M., Hernan, M. A., and Brumback, B. (2000), “Marginal structural models and causal inference in epidemiology,” Epidemiology, 550–560.

Robins, J. M. and Richardson, T. S. (2010), “Alternative graphical causal models and the identification of direct effects,” Causality and psychopathology: Finding the determinants of disorders and their cures, 84, 103–158.

Robins, J. M., Richardson, T. S., and Shpitser, I. (2022), “An interventionist approach to mediation analysis,” in Probabilistic and causal inference: the works of Judea Pearl, pp. 713–764.

Rosenbaum, J. F. (2024), “Functional Unblinding in Pivotal Studies and the Future of Psychedelic Medicine,” The Journal of Clinical Psychiatry, 85, 56467.

Schenberg, E. E. (2025), “From Efficacy to Effectiveness: Evaluating Psychedelic Randomized Controlled Trials for Trustworthy Evidence-Based Policy and Practice,” Pharmacology Research & Perspectives, 13, e70097.

Stensrud, M. J., Nevo, D., and Obolski, U. (2024), “Distinguishing Immunologic and Behavioral Effects of Vaccination,” Epidemiology, 35, 154–163.

Stensrud, M. J., Robins, J. M., Sarvet, A., Tchetgen Tchetgen, E. J., and Young, J. G. (2023), “Conditional separable effects,” Journal of the American Statistical Association, 118, 2671– 2683.

Stensrud, M. J., Young, J. G., Didelez, V., Robins, J. M., and Hernán, M. A. (2022), “Separable Effects for Causal Inference in the Presence of Competing Events,” Journal of the American Statistical Association, 117, 175–183.

Szigeti, B., Bradley, E., and Woolley, J. (2024a), “Unmasking bias and MDMA-assisted therapy,”.

Szigeti, B. and Heifets, B. D. (2024), “Expectancy Effects in Psychedelic Trials,” Biological Psychiatry: Cognitive Neuroscience and Neuroimaging, 9, 512–521, therapeutic Tripping: Mechanisms Underlying the Clinical Potential of Psychedelics.

Szigeti, B., Nutt, D., Carhart-Harris, R., and Erritzoe, D. (2023), “The difference between ‘placebo group’and ‘placebo control’: A case study in psychedelic microdosing,” Scientific Reports, 13, 12107.

Szigeti, B., Weiss, B., Rosas, F. E., Erritzoe, D., Nutt, D., and Carhart-Harris, R. (2024b), “Assessing expectancy and suggestibility in a trial of escitalopram v. psilocybin for depression,” Psychological Medicine, 54, 1717–1724.

Tambling, R. B. (2012), “A literature review of therapeutic expectancy effects,” Contemporary Family Therapy, 34, 402–415.

Tchetgen Tchetgen, E. J., Ying, A., Cui, Y., Shi, X., and Miao, W. (2024), “An introduction to proximal causal inference,” Statistical Science, 39, 375–390.

U.S. Food and Drug Administration (2024), “Complete Response Letter – NDA 215455 (Ref. ID 5427030),” Retrieved September 5, 2025, from Psychedelic Alpha.

van Elk, M., Fejer, G., Lempe, P., Prochazckova, L., Kuchar, M., Hajkova, K., and Marschall, J. (2022), “Effects of psilocybin microdosing on awe and aesthetic experiences: a preregistered field and lab-based study,” Psychopharmacology, 239, 1705–1720.

VanderWeele, T. J. and Hernan, M. A. (2013), “Causal inference under multiple versions of treatment,” Journal of causal inference, 1, 1–20.

VanderWeele, T. J., Vansteelandt, S., and Robins, J. M. (2014), “Effect decomposition in the presence of an exposure-induced mediator-outcome confounder,” Epidemiology, 25, 300–306.

Vollenweider, F. X., Vollenweider-Scherpenhuyzen, M. F., Bäbler, A., Vogel, H., and Hell, D. (1998), “Psilocybin induces schizophrenia-like psychosis in humans via a serotonin-2 agonist action,” Neuroreport, 9, 3897–3902.

Vuong, Q., Metcalfe, R. K., Ling, A., Ackerman, B., Inoue, K., and Park, J. J. (2025), “Systematic review of applied transportability and generalizability analyses: A landscape analysis,” Annals of Epidemiology.

Wen, J., Satyanarayanan, S. K., Li, A., Yan, L., Zhao, Z., Yuan, Q., Su, K.-P., and Su, H. (2024), “Unraveling the impact of Omega-3 polyunsaturated fatty acids on blood-brain barrier (BBB) integrity and glymphatic function,” Brain, Behavior, and Immunity, 115, 335–355.

Zhang, Z., Kotz, R. M., Wang, C., Ruan, S., and Ho, M. (2013), “A causal model for joint evaluation of placebo and treatment-specific effects in clinical trials,” Biometrics, 69, 318–327.

For use of this code or vignette, please cite the following papers:

